# The Contributions of Rare Inherited and Polygenic Risk to ASD in Multiplex Families

**DOI:** 10.1101/2022.04.05.22273459

**Authors:** Timothy S Chang, Matilde Cirnigliaro, Stephanie A Arteaga, Laura Pérez-Cano, Elizabeth K Ruzzo, Aaron Gordon, Lucy Bicks, Jae-Yoon Jung, Jennifer K Lowe, Dennis P Wall, Daniel H Geschwind

## Abstract

Autism Spectrum Disorder (ASD) has a complex genetic architecture involving contributions from *de novo* and inherited variation. Few studies have been designed to address the role of rare inherited variation, or its interaction with polygenic risk in ASD. Here, we performed whole genome sequencing of the largest cohort of multiplex families to date, consisting of 4,551 individuals in 1,004 families having 2 or more affected children with ASD. Using this study design, we identify seven novel risk genes supported primarily by rare inherited variation, finding support for a total of 74 genes in our cohort and a total of 152 genes after combining with other studies. Probands demonstrated an increased burden of mutations in 2 or more known risk genes (KARGs) — in three families both probands inherited protein truncating variants in two KARGs. We also find that polygenic risk is over transmitted from unaffected parents to affected children with rare inherited variants, consistent with combinatorial effects in the offspring, which may explain the reduced penetrance of these rare variants in parents. We also observe that in addition to social dysfunction, language delay is associated with ASD polygenic risk over-transmission. These results are consistent with an additive complex genetic risk architecture of ASD involving rare and common variation and further suggest that language delay is a core biological feature of ASD.

## Introduction

The last decade has seen enormous progress in identifying genes imparting risk for autism spectrum disorder (ASD) (O’Roak et al. 2012; Iossifov et al. 2014; O’Roak et al. 2014; De Rubeis et al. 2014; Krumm et al. 2015; Sanders et al. 2015; Turner et al. 2016; Stessman et al. 2017; Ruzzo et al. 2019; Satterstrom et al. 2020), identifying over 100 genes whose risk is due primarily to rare *de novo* copy number variants or protein truncating variants (Yuen et al. 2017; Werling et al. 2018; Feliciano et al. 2019; Satterstrom et al. 2020; Sanders et al. 2015; Turner et al. 2017; Dong et al. 2014; Iossifov et al. 2014; O’Roak et al. 2012; Neale et al. 2012; Sebat et al. 2007; Marshall et al. 2008; Bucan et al. 2009). The majority of these risk genes appear to act pleiotropically, contributing to a broad range of neurodevelopmental disorders in addition to ASD, ranging from epileptic encephalopathy, attention-deficit/hyperactivity disorder, intellectual disability, and schizophrenia in some cases (Epi4K Consortium et al. 2013; Satterstrom et al. 2019; Stessman et al. 2017; Satterstrom et al. 2020; Singh et al. 2017; Cross-Disorder Group of the Psychiatric Genomics Consortium. 2019). These findings have delivered on the original promise of genetic investigations by galvanizing functional genomic and neurobiological studies that have begun to yield mechanistic insight into ASD pathophysiology (de la Torre-Ubieta et al. 2016; Geschwind and State 2015; Abrahams and Geschwind 2008).

However, ASD is also highly heritable (Gaugler et al. 2014; Colvert et al. 2015; Sandin et al. 2017; Bai et al. 2019) and therefore is expected to have a substantial contribution from common (Gaugler et al. 2014; Weiner et al. 2017) and rare variation transmitted from parents to their affected offspring. Indeed, at least 50% of genetic risk is predicted to be due to common variation, 15-20% is due to *de novo* variation and other Mendelian forms, and the remaining genetic risk is yet to be determined (Grove et al. 2019; Gaugler et al. 2014; Iossifov et al. 2014; De Rubeis et al. 2014; Satterstrom et al. 2020). Even with substantial sized cohorts (greater than 25,000), genome wide association studies (GWAS) in ASD have to date only identified five loci reaching genome wide significance (Grove et al. 2019). This is consistent with high genetic heterogeneity, as well as contribution from other rare forms of inherited variation.

Another potential contribution to the preponderance of genes supported by *de novo* variation, and less success in detecting inherited variation, may be study designs with a heavy focus on families with simplex ASD, having only a single proband and no family history, so that probands are depleted for inherited genetic risk for ASD (Ronemus et al. 2014; Leppa et al. 2016; Yuen et al. 2015; Feliciano et al. 2019). Indeed, our recent study of families having 2 or more affected children, demonstrated a different genetic architecture in these multiplex ASD families (Lajonchere and AGRE Consortium 2010; Ruzzo et al. 2019) with a significant signal from rare, inherited variants and a less significant contribution from *de novo* variation (Ruzzo et al. 2019). Studying multiplex families allowed us to identify 16 new ASD genes supported predominantly by inherited rare variants in addition to *de novo* variants (Ruzzo et al. 2019).

In this current study, we performed whole genome sequencing on the largest collection of ASD multiplex families to date, including 4,551 individuals, comprised of 1,836 probands and 418 unaffected siblings from 1,004 families to further explore the contribution of rare and common inherited variation to ASD genetic risk architecture. We identified 74 ASD risk genes harboring rare protein disrupting or damaging missense variants, seven of which were not identified in previous studies and thus are novel risk genes. We also leverage WGS to assess the contribution of polygenic risk (PGR) in those with inherited and *de novo* variants to further frame the genetic architecture of ASD. We show that while PGR is not over-transmitted to ASD probands harboring *de novo* variants, PGR is significantly over-transmitted to those harboring inherited protein coding rare variants and those with a more severe ASD phenotype. These data provide new evidence supporting a complex view of ASD genetic architecture that includes a mix of common and rare genetic variation, whereby PGR contributes to ASD risk in combination with rare inherited protein truncating variants. The contribution of PGR to autism risk may also explain why parents harboring rare protein disrupting variants are asymptomatic, since they lack the polygenic contribution from common variants that is co-transmitted along with rare risk variants to their affected children.

## Results

We performed whole genome sequencing at a mean >30x coverage in 4,551 individuals from 1,004 multiplex ASD families (Table S1). Study individuals were from the Hartwell Autism Research and Technology Initiative (iHART) cohort within the Autism Genetic Resource Exchange (AGRE) (Lajonchere and AGRE Consortium 2010), and included an initial smaller published subset of this cohort (493 multiplex ASD families, (Ruzzo et al. 2019)). This analysis of multiplex ASD families included 1,836 affected children and 418 unaffected children where both biological parents were sequenced (fully-phaseable) (Figure 1) and is more than double in size compared to the previous study (Ruzzo et al. 2019). Cases were diagnosed according to widely used protocols that have been well validated (Lord, Rutter, and Le Couteur 1994; Lord et al. 2000), and details of clinical ascertainment in AGRE have been previously published (Leppa et al. 2016; Geschwind et al. 2001).

**Figure 1.**
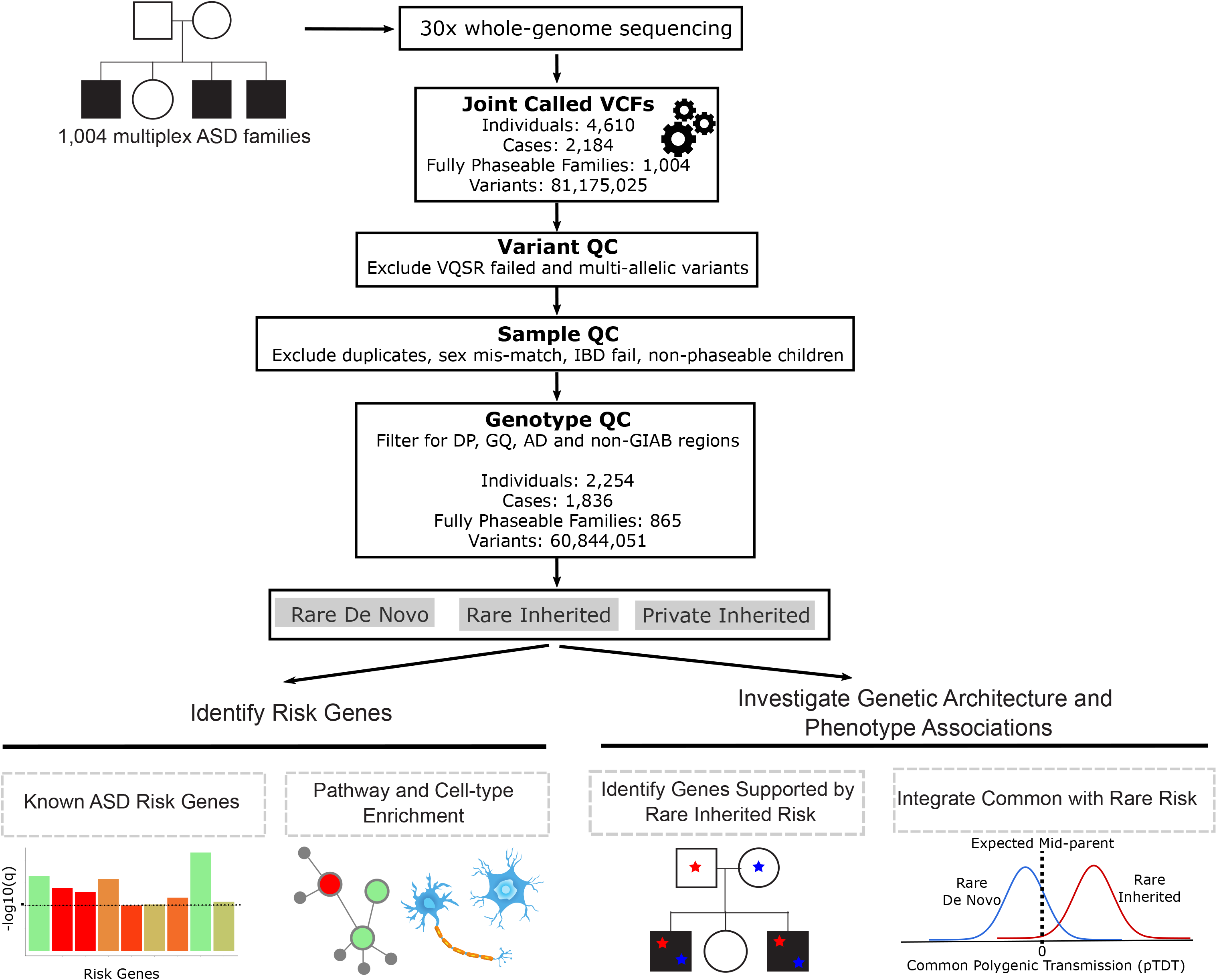
Overview diagram of study analyses. Whole genome sequencing was performed on 1,004 multiplex ASD variants. Single nucleotide variants (SNV) and small insertions/deletions (Indels) were called using the Genome Analysis Toolkit (GATK). Variant, sample and genotype quality control was performed. Rare *de novo*, rare inherited, and private inherited variants were included for downstream analyses. We identified ASD risk genes using the Transmission and *de novo* association (TADA) analysis. Risk genes were characterized using integrative genomics. We also investigated the genetic architecture of ASD phenotypes. We identified genes supported by rare inherited risk by evaluating the phenotype of children with multiple inherited risk genes. We tested if common ASD polygenic risk was over transmitted to affected versus unaffected children stratified by rare *de novo* and inherited variants.

Of the 1,004 multiplex ASD families, 714 families have 2 fully-phaseable affected children (71.1%), 109 families have 3 fully-phaseable affected children (10.8%), 8 families have 4 fully-phaseable affected children (0.8%), 4 families have 5 fully-phaseable affected children (0.4%), and 368 families have at least 1 fully-phaseable unaffected child (36.7%). Among the fully-phaseable children, 1620 are males and 634 are females. We also utilized 76 families with monozygotic concordant pairs to inform quality control (Ruzzo et al. 2019).

### Overall genetic architecture and transmission patterns in multiplex families

Whole genome sequencing yielded an average coverage of 37.48X with 80.6% of bases covered at ≥30X and 99.2% of bases covered at ≥10X (Figures S1A-S1E). Quality metrics were within the expected range for whole genome sequencing (WGS) (J. Wang et al. 2015) including a transition to transversion ratio of 2.155, a heterozygous to non-reference homozygous ratio of 1.6, and dbSNP concordant rate of 99.92%. Our WGS processing pipeline retained high quality variants, for example by removing variant calls for samples with a genotype quality (GQ)<25, depth (DP)<10 and alternate allele depth to depth (ALT AD/DP)<0.2 (Methods, Figure S2). In addition, we applied a validated machine learning approach based on random forests, Artifact Removal by Classifier (ARC) (Ruzzo et al. 2019), to distinguish true rare *de novo* variants (RDNVs) from likely sequencing, mapping errors or artifacts from lymphoblastoid culture (Methods) (Conrad et al. 2011; 1000 Genomes Project Consortium et al. 2015).

After ARC, we find an overall rate of 53 rare *de novo* variants per subject, with no differences between cases and controls (Methods, Wilcoxon rank sum test p=0.20), consistent with published literature (Besenbacher et al., 2016; Conrad et al., 2011; Kong et al., 2012; Michaelson et al., 2012; Turner et al., 2016; Yuen et al., 2017). As previously, we find a robust association of paternal age and *de novo* association rate (Figure S3, Methods, 1.02 RDNVs per year of paternal age, p<2.2 x 10^-16^) (Deciphering Developmental Disorders Study 2017; Francioli et al. 2015; Goldmann et al. 2016; Michaelson et al. 2012). The mean number of rare inherited variants (MAF<0.001, see Methods for additional quality criteria), private inherited variants, and rare *de novo* variants (protein truncating, missense or synonymous) per subject was 145.5, 32.4, and 0.46 respectively. Consistent with the hypothesized and previously observed depletion of rare *de novo* ASD risk in multiplex families (Leppa et al. 2016; Ruzzo et al. 2019), we observed no enrichment of rare *de novo* PTVs (logistic regression, p = 0.79) and missense variants (logistic regression, p = 0.46) in affected children compared to unaffected children (Figure 2A). We obtained similar results when comparing the rates of these variant classes in highly constrained genes (pLI≥0.9, pLI≥0.995) between affected and unaffected children (logistic regression, pLI≥0.9 missense p = 0.63, pLI≥0.9 PTV p = 0.69, pLI≥0.995 missense p = 0.07, pLI≥0.995 PTV p = 0.99) (Figures 2B and 2C). We next assessed the rate of rare inherited variants in affected and unaffected children. We found no excess of rare inherited protein truncating variants (PTVs) (Methods, logistic regression, p = 0.60) and missense variants (logistic regression, Polyphen-2 mis3 (Adzhubei et al., 2010) p = 0.62; MPC ≥ 1 (Samocha et al. 2017) p=0.77) in affected children (Figure 2D). The same held true when limiting the analysis to rare inherited PTV and missense variants in highly constrained genes (logistic regression, pLI≥0.9 missense mis3 p = 0.78, pLI≥0.9 missense MPC≥1 p=0.4, pLI≥0.9 PTV p = 0.12, pLI≥0.995 missense mis3 p = 0.96, pLI≥0.995 missense MPC≥1 p = 0.22, pLI≥0.995 PTV p = 0.89) (Figures 2E and 2F). Similarly, we observed no difference in the rates of private inherited variants (overall, PTV, and missense variant rates in all genes and in highly constrained genes) between affected and unaffected children (Figures 2G-2I).

**Figure 2.**
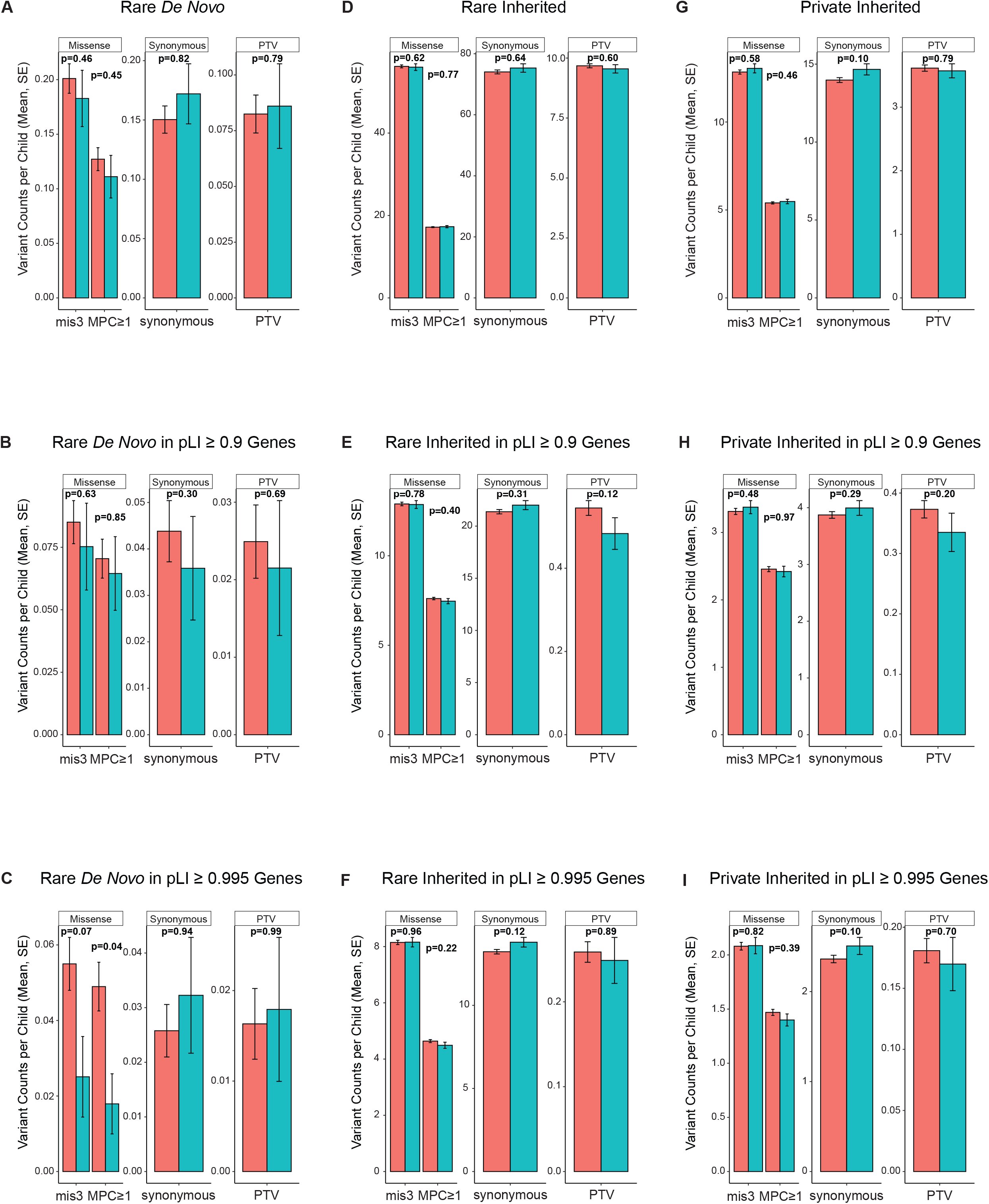
(A-C) Rate of rare *de novo* variants in all genes, genes with pLI≥0.9, and genes with pLI>0.995 stratified by variant type. (D-F) Rate of rare inherited variants in all genes, genes with pLI>0.9, and genes with pLI≥0.995 stratified by variant type. (G-I) Rate of private inherited variants in all genes, genes with pLI≥0.9, and genes with pLI>0.995 stratified by variant type. mis3 = Polyphen-2 missense 3, MPC≥1=missense deleterious metric

### Non-coding variation in multiplex families

We extended our investigation of the distinct genetic architecture of ASD multiplex families to non-coding regions of the genome. Previous category-wide association studies (CWAS) carried out in simplex families have suggested a biologically plausible association of promoter regions with ASD supported by *de novo* variation (Werling et al. 2018; An et al. 2018). Specifically, it has been observed that this non-coding association signal is driven by evolutionary conservation and that it is stronger in promoter regions of conserved loci more distal to the transcription start site (TSS) (An et al. 2018). Therefore, we first compared non-coding variant burden between affected and unaffected children in iHART for variants mapping to 3kb (2kb upstream and 1kb downstream of the TSS) promoters of all genes. We found no significant enrichment of promoter rare inherited (logistic regression, p = 0.81), private inherited (logistic regression, p = 0.14), and rare *de novo* (logistic regression, p = 0.5) variants in cases (Figures S4A, S4B, and S4C). These results held true when focusing the rare and private inherited non-coding variant testing to promoters of ASD risk genes (Methods, Figures S4D and S4E). We extended the promoter definition from 2kb to 10kb upstream of the transcription start site with the aim of capturing distal promoter regions. The extension of promoter regions to 11kb (including 1kb downstream of TSS) did not alter the previous results (logistic regression; rare inherited, p = 0.87; private inherited, p = 0.11; rare *de novo*, p = 0.55) (Figures S4A, S4B, and S4C).

The strongest enrichment of *de novo* conserved promoter variants in ASD simplex cases in prior work has been found at transcription factor binding sites (TFBSs) (An et al. 2018). Hence, we filtered variants in the 11kb promoter regions of all genes by keeping variants in regions defined by three different genome-wide maps of TFBSs: (1) cis-regulatory modules (CRMs), which are clusters of predicted TFBSs (Gheorghe et al. 2019), as well as (2) cistromes and (3) cismotifs, which are experimentally and technically reproducible TF binding regions (Vorontsov et al. 2018). Again, we found no significant excess of TFBS rare inherited (logistic regression; CRMs, p = 0.99; cistromes, p = 0.91; cismotifs, p = 0.9), private inherited (logistic regression; CRMs, p = 0.21; cistromes, p = 0.13; cismotifs, p = 0.15), and rare *de novo* (logistic regression; CRMs, p = 0.98; cistromes, p = 0.58; cismotifs, p = 0.76) variants in iHART affected children compared to unaffected children (Figures S4F, S4G, and S4H).

Finally, we mapped non-coding variants within the 11kb promoters of all genes and non-coding variants across the whole genome to the brain-specific regulatory regions under sequence constraint within the human lineage, identified through the recent co-localization approaches (Xu et al. 2020). Xu et al. previously showed these brain-specific regulatory regions were enriched for non-coding *de novo* variants in affected compared to unaffected children from ASD simplex families (Xu et al. 2020). In our study, we found no significant burden of brain-specific constrained functional non-coding rare inherited (logistic regression; 11kb promoter intersected set, p = 0.93; genome intersected set, p = 0.82), private inherited (logistic regression; 11kb promoter intersected set, p = 0.27; genome intersected set, p = 0.15), and rare *de novo* (logistic regression; 11kb promoter intersected set, p = 0.54; genome intersected set, p = 0.18) variants in iHART cases (Figures S4I, S4J, and S4K). We were not able to replicate in iHART known signals for non-coding *de novo* variant sets (Table S2), consistent with findings from our previous study (Ruzzo et al. 2019). This could be the result of the *de novo* depletion observed in this multiplex cohort and decreased power for non-coding association testing.

Previous power calculation for non-coding variation across ranges of relative risks, category sizes, and sample sizes assessed with the CWAS framework in simplex families confirmed how different the frequency and effect size of non-coding variant categories are from the well-defined and functionally characterized coding variants (Werling et al. 2018). It was estimated that over 8,000 simplex families would be needed to have sufficient power to discover a *de novo* non-coding class equivalent to *de novo* missense variation (Werling et al. 2018). From the filtering strategies we performed, we narrowed iHART non-coding variants to putatively functional sets more likely to be associated with ASD. Across all tested sets we found very small effect sizes for non-coding variation (Table S2), which results in low power at our current sample size and is consistent with previous non-coding investigation focusing on *de novo* variants (Werling et al. 2018; An et al. 2018) (Table S3). In addition, taking advantage of the multiplex family study design, we assessed the range of effect sizes for non-coding rare inherited variants, confirming estimates similar to those for the non-coding *de novo* variants (Werling et al. 2018) (Table S2). Based on the observed effect sizes from our study, we computed power by simulation (Table S3) and predicted that a sample size of at least 8,000 fully-phaseable children would provide sufficient power for the investigation of private inherited variants in 11kb promoters of all genes, whereas 9,000 children would be needed to test association of non-coding private inherited variants in functional regions such as cistromes, cismotifs, and brain-specific co-localized regions (Figure S4L-N).

### Seven New ASD Risk Genes

We do not observe a global excess for damaging rare *de novo* variants in affected children in these multiplex families, consistent with a lower contribution and distinct genetic architecture compared with simplex families. However, we still do identify pathogenic *de novo* variants in previously established ASD-risk genes in some affected family members. We therefore used the Transmitted And *De novo* Association (TADA) test (He et al. 2013), a Bayesian framework that combines inherited and *de novo* genetic variants, to identify ASD risk genes in this cohort. Qualifying variants included rare *de novo* or transmitted PTVs and *de novo* missense3 variants predicted to damage the encoded protein (Adzhubei et al., 2010) (Methods, Figure 3A, and Table S4). We combined this extended multiplex cohort with 1,660 affected children and their fully-phaseable parents with qualifying variants from previous ASD genetic analyses (Sanders et al. 2015; Iossifov et al. 2014; De Rubeis et al. 2014; Pinto et al. 2014) (Methods, Figure 3A, Figure S5A). We identified 74 genes significantly associated with ASD (FDR<0.1) (Figure 3B). Of these 74 genes, 46% were supported only by *de novo* variants. The remaining genes (54%) were supported by inherited variants, half of which had more than 50% of genetic variants with an inherited component (Figure 3B). There were nine genes identified in this study, but not our previous iHART study (Ruzzo et al. 2019). Two genes (*MED13L, KMT2A*) were previously identified in ASD studies and harbored *de novo* variants (Simons Foundation Autism Research Initiative 2020; Satterstrom et al. 2020) as was also shown in our study. Seven genes are novel, having not been identified in previous studies (*PLEKHA8, PRR25, FBXL13, VPS54, SLFN5, SNCAIP, TGM1*) (Figure 3C). Consistent with the distinct architecture involving more inherited variation in multiplex ASD, five of these genes were predominantly supported by inherited protein truncating variants (*PRR25, FBXL13, VPS54, SLFN5, TGM1*).

**Figure 3.**
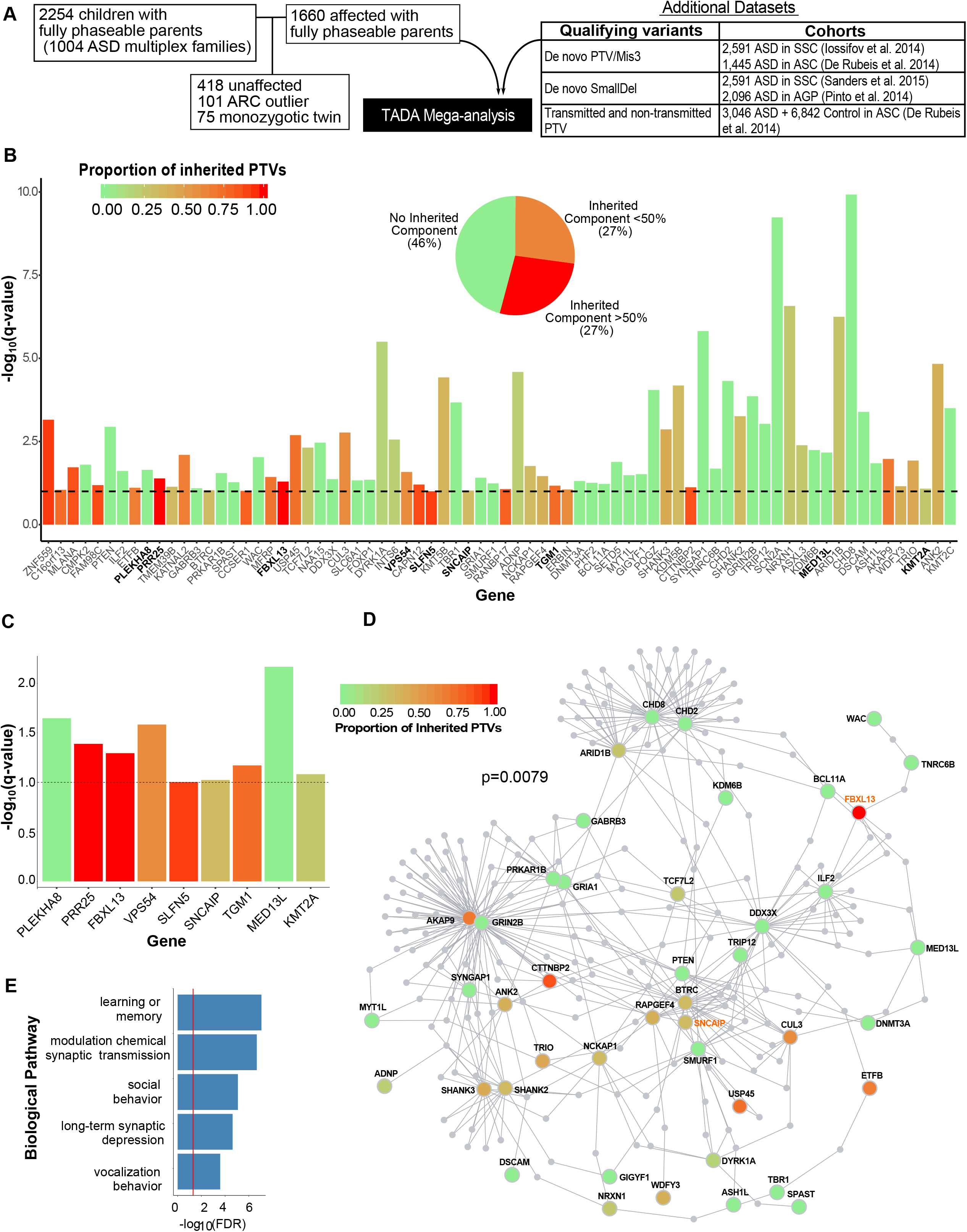
74 ASD risk genes identified by Transmission and *de novo* association (TADA) analysis. (A) Overview of iHART subjects and additional datasets included in the TADA analysis. (B) 74 ASD risk genes identified in the iHART TADA analysis (FDR<0.1) are displayed as the -log10(q-value). The nine newly identified ASD risk genes from our expanded iHART cohort are bold. The dashed horizontal line marks the FDR=0.1 threshold. Bars are colored by the proportion of inherited protein truncating variants (PTVs) for each gene (inherited PTVs/(inherited PTVs + *de novo* PTVs + *de novo* Mis3 + *de novo* small deletions). (C) The nine newly identified ASD risk genes from our expanded iHART cohort. Bars are colored according to (B). (D) Protein-protein interaction network formed by the 74 ASD risk genes using Disease Association Protein-Protein Link Evaluator (DAPPLE). Genes are colored by the proportion of inherited PTVs. The gene name for two of the seven newly identified ASD risk genes are in orange. (E) Gene Ontology enrichment terms for 39 significant seed genes from the protein-protein interaction network

We next assessed the shared biological functions among the 74 ASD risk genes from our TADA analysis. These genes formed a significant indirect protein-protein interaction (PPI) network (Methods, p=0.0079) involving 39 genes as significant seed genes, meaning they were more connected to other genes in the network than expected by chance (Figure 3D). These genes enriched for gene ontology terms including synaptic transmission, learning, social behavior and long-term synaptic depression (Figure 3E).

Previous work has demonstrated that ASD risk genes are most highly expressed as a group during brain development (Satterstrom et al. 2020; Ruzzo et al. 2019; Sanders et al. 2015; De Rubeis et al. 2014; Iossifov et al. 2014; Krumm et al. 2015). We subsequently used human brain fetal single cell transcriptomics (Polioudakis et al. 2019) to determine in which cell types the 74 ASD risk genes were expressed. We observed enriched expression of ASD risk genes in excitatory neurons (Expression Weighted Cell Type Enrichment (EWCE) migrating excitatory neuron FDR=0.0015, maturing excitatory neuron FDR=0.00016, Methods, Figures 4A,4B) and interneurons (EWCE interneurons CGE FDR=0.027, interneuron MGE FDR=0.04, Methods, Figures 4A,4B) as expected based on previous results (Ruzzo et al. 2019; Satterstrom et al. 2019)

**Figure 4.**
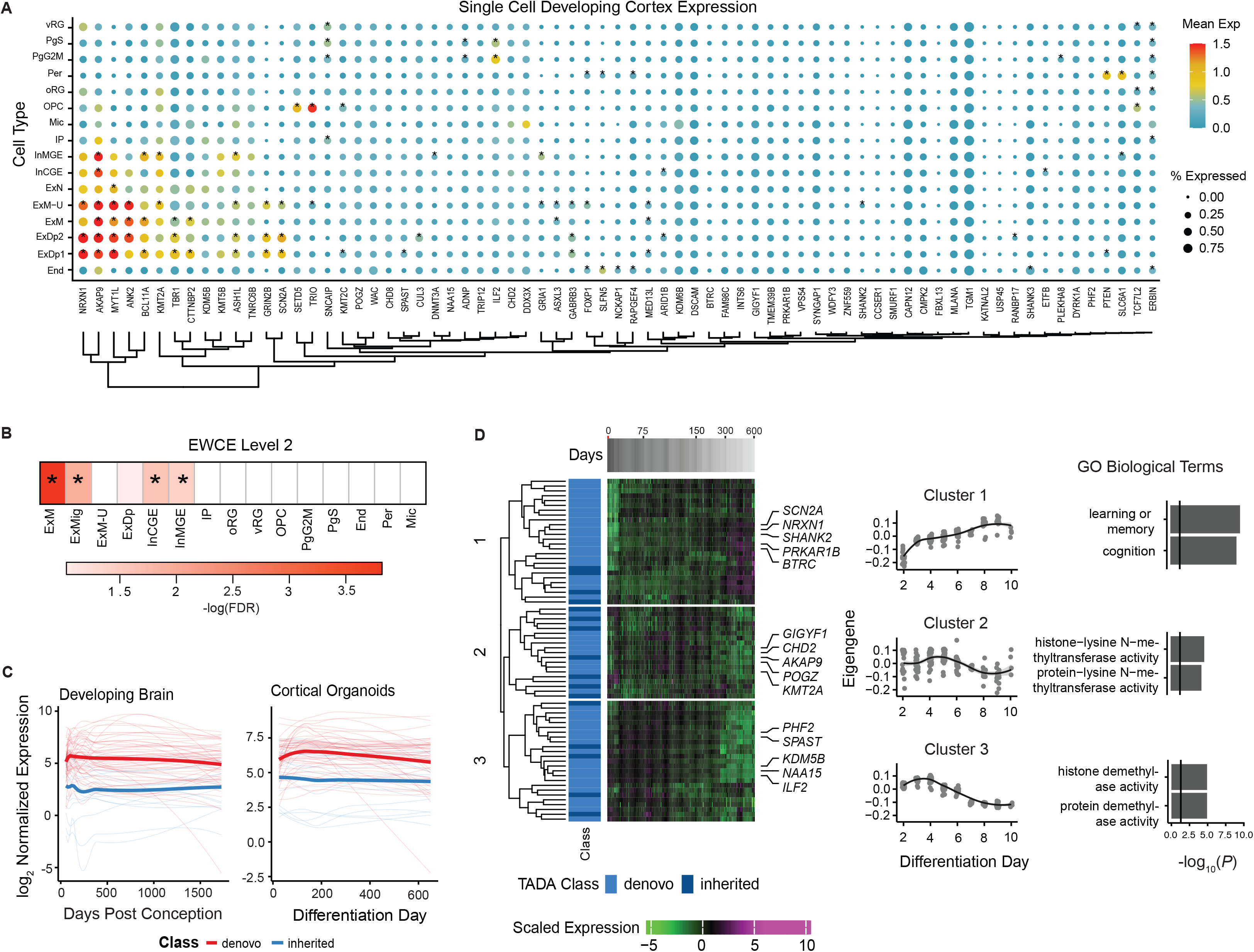
Characterization of TADA ASD risk genes during development. (A) Mean expression and percentage of cells expressing TADA ASD risk genes from single cell data of the human fetal brain cortex (Polioudakis et al. 2019). * Indicates cell type specific TADA ASD risk genes (Methods). (B) Cell types enriched in TADA ASD risk genes tested using Expression Weighted Cell Type Enrichment (EWCE). * FDR<0.05. (C) TADA ASD risk genes variation during development using a long-term maturation human cortical spheroid (hCS) and BrainSpan. ASD risk genes are stratified by those with predominantly inherited or *de novo* support. (D) Clustering of the ASD risk genes from hCS trajectory data identified 3 clusters. Days post conception, TADA class (predominant inherited or *de novo*) and scaled expression are shown. The average expression of each cluster and significant gene ontology biological terms are shown. vRG= ventral radial glia, PgS = cycling progenitor (S phase), PgG2M= cycling progenitor (G2M phase), Per=pericyte, oRG=outer radial glia, OPC=oligodendrocyte precursor, Mic=microglia, IP=intermediate progenitor, InMGE=Interneuron medial ganglionic eminence, InCGE=interneuron caudal ganglionic eminence, ExN=excitatory neuron, ExM−U=maturing excitatory neuron upper enriched, ExM=maturing excitatory neuron, ExDp2=excitatory neuron deep layer 2, ExDp1= excitatory neuron deep layer 1, End=endothelial, ExMig = migrating excitatory neuron, ExDp = excitatory neuron deep

We next evaluated how these 74 ASD risk genes varied during development using data from two relevant systems: long-term maturation in 3-dimensional induced pluripotent stem cell (IPSC)-derived human forebrain organoid model, human cortical spheroids (hCS), which have been shown to largely recapitulate human fetal brain development up to 600 days postnatally in vitro (Gordon et al. 2021) and the BrainSpan dataset, which represents an *in vivo* reference for cortical development (Stein et al. 2014; Kang et al. 2011; M. Li et al. 2018). Given the developmental function of ASD risk genes identified through *de novo* variants (Ronemus et al. 2014; Satterstrom et al. 2020; Iossifov et al. 2014), genes with predominantly *de novo* variant support (<50% inherited component support) were mostly highly expressed early in development in both the hCS model and BrainSpan (Figure 4C), consistent with earlier findings (Parikshak, Gandal, and Geschwind 2015). Interesting, the average expression of genes with predominantly *de novo* variant support had higher expression compared to the average expression of genes with predominantly inherited variant support during this period as well.

We also clustered the ASD risk genes using the hCS trajectory data, which identified three clusters. ASD risk genes with greater than a 50% inherited component were represented in all clusters. Cluster 1 included genes which gradually rose during development and were enriched for genes involved in learning and memory (Figure 4D). Clusters 2 and 3 included genes that were elevated early in development and were enriched for genes with histone transferase activity (Figure 4D). Overall, these results show ASD risk genes with predominant *de novo* support have highest expression early in development and decline over time, while a subset of ASD risk genes with both *de novo* or inherited support (Cluster 1) that are involved in cognition and learning rise more gradually across fetal neuronal development.

### Known ASD Risk Genes in Multiplex Families support Oligogenic Burden

We next collated a list of all known ASD risk genes to determine their contribution to risk in multiplex families by identifying unique ASD risk genes from our study and recent large ASD genetic studies (Satterstrom et al. 2020; Sanders et al. 2015) with an FDR < 0.1, yielding a total of 152 genes (Table S5). Not surprisingly, affected children had significantly more rare inherited protein truncating variants in known ASD risk genes (KARGs) compared to unaffected children (p=0.0196, Figure 5A). In contrast to observations in simplex families (O’Roak et al. 2012; Iossifov et al. 2014; O’Roak et al. 2014; De Rubeis et al. 2014; Krumm et al. 2015; Sanders et al. 2015; Turner et al. 2016; Stessman et al. 2017; Satterstrom et al. 2020), rare *de novo* protein truncating variants in known ASD risk genes were not increased in affected versus unaffected children (p=0.366, Figure 5B).

**Figure 5.**
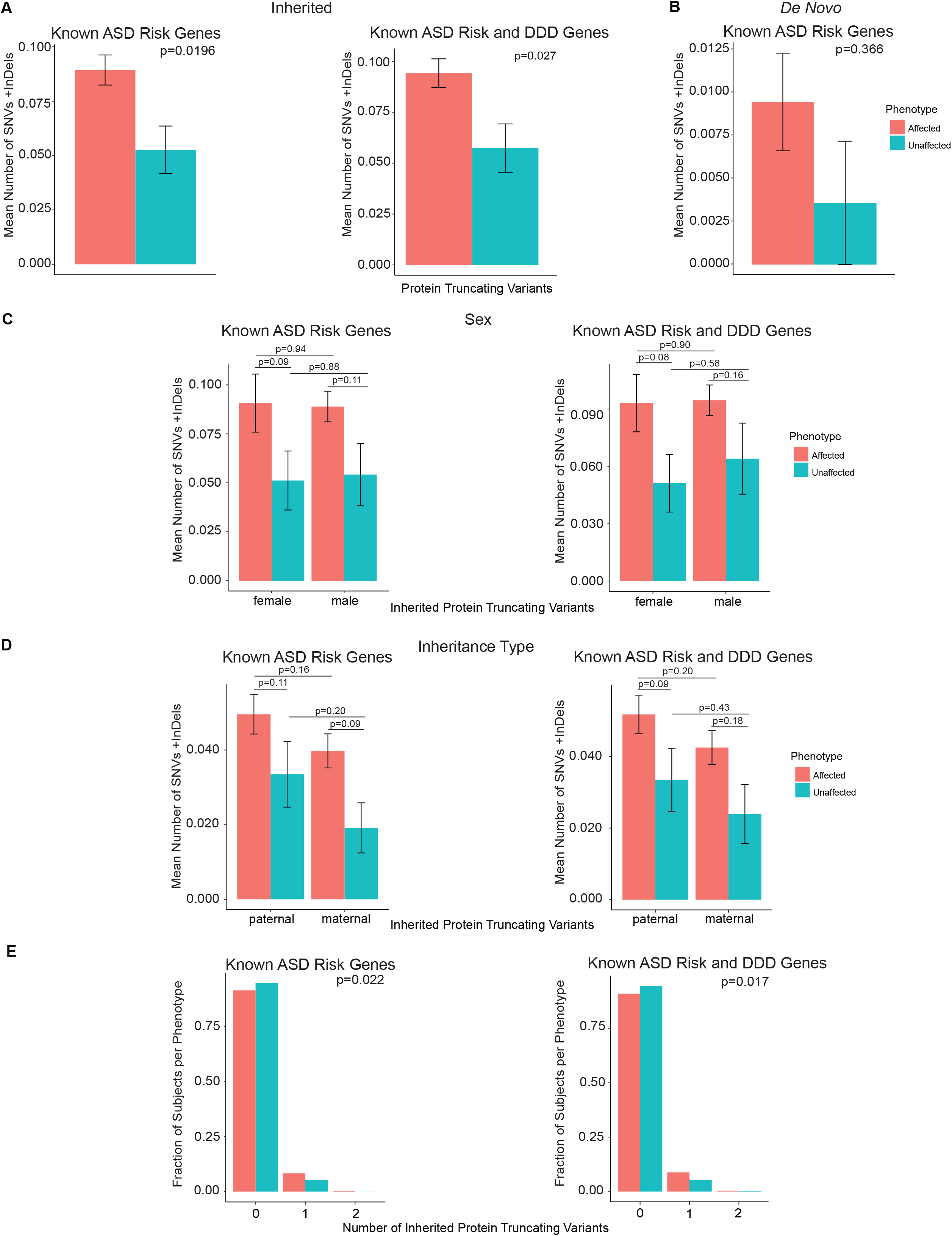
Known ASD risk genes variant rates, and known ASD and developmental disorder risk genes variant rate. Mean number of protein truncating variants in affected versus unaffected children with inherited (A) and *de novo* (B) variants. Mean number of inherited protein truncating variants in affected versus unaffected children stratified by sex (C) and inheritance type (D). Fraction of subjects and number of genes with protein truncating variants from known ASD risk genes or known ASD and developmental disorder risk genes (E).

Given the lower female to male ratio of ASD and multiple threshold theory of a female protective effect (Maenner et al. 2020; Fombonne 2009; Werling and Geschwind 2013), we examined rare inherited protein truncating variants in known ASD risk genes by sex. There was no difference in mean number of rare inherited protein truncating variants in known ASD risk genes in affected females versus affected males (logistic regression, p=0.94, Figure 5C, Methods), or affected versus unaffected children stratified by sex (Figure 5C). Both paternal and maternal inheritance of variants has been associated with ASD risk (Brandler et al. 2018; Jacquemont et al. 2014). We observe no difference between the two modes of inheritance in this cohort (Figure 5D), although we acknowledge that larger population scale studies may be needed to see more subtle differences.

We next examined whether there were affected children who inherited more than one PTV in a known ASD risk gene (KARG), referred to as oligogenic transmission (Turner et al. 2017). Indeed, we found that there was an excess of affected children with one or two protein truncating variants in KARGs (Mann-Whitney test, p=0.022, Figure 5E, Methods). But, we did not observe this effect when considering mutation burden across all genes, consistent with the recognized mutation intolerance of KARGs (p=0.486, Figure S5B) (Sanders et al. 2015; Samocha et al. 2017). We also evaluated known risk genes for ASD and developmental delay (DD) (Deciphering Developmental Disorders Study 2017), which totaled 208 genes (Table S5). Similar to observations in known ASD risk genes, the ASD and DD risk genes showed an increase in the number of inherited protein truncating variants (p=0.027, Figure 5A) and an excess of one or two protein truncating variants (p=0.017, Figure 5E) in affected children. There was no significant difference of known risk genes for ASD and developmental delay by sex (Figure 5C) or inheritance type (Figure 5D).

To further investigate these observations, we identified six affected children carrying two rare inherited PTVs in known ASD risk genes: specifically, these children are sibling pairs from three different families (Figure 6). In family A, the two affected siblings inherit rare PTVs in both *CAPN12* and *FBXL13* from the father, while the unaffected child has inherited only the *FBXL13* variant. In family B, both affected children inherited a PTV in *MFRP* from the father and a PTV in *SHANK3* from the mother. Interestingly, in family C, of the three affected dizygotic twins, the two females inherited PTVs in both *RANBP17* and *WDFY3* from the father, while the male did not. Intriguingly, the two female affected twins had their first steps and phrases much later than their male affected twin who, in contrast to the two affected females, did not show severely impaired social behavior and social interaction skills. Of note, RNA-seq analysis on 120 human fetal brains highlighted *RANBP17* as a high-confidence ASD risk gene exhibiting higher expression in females (FDR < 0.1) (O’Brien et al. 2019).

**Figure 6.**
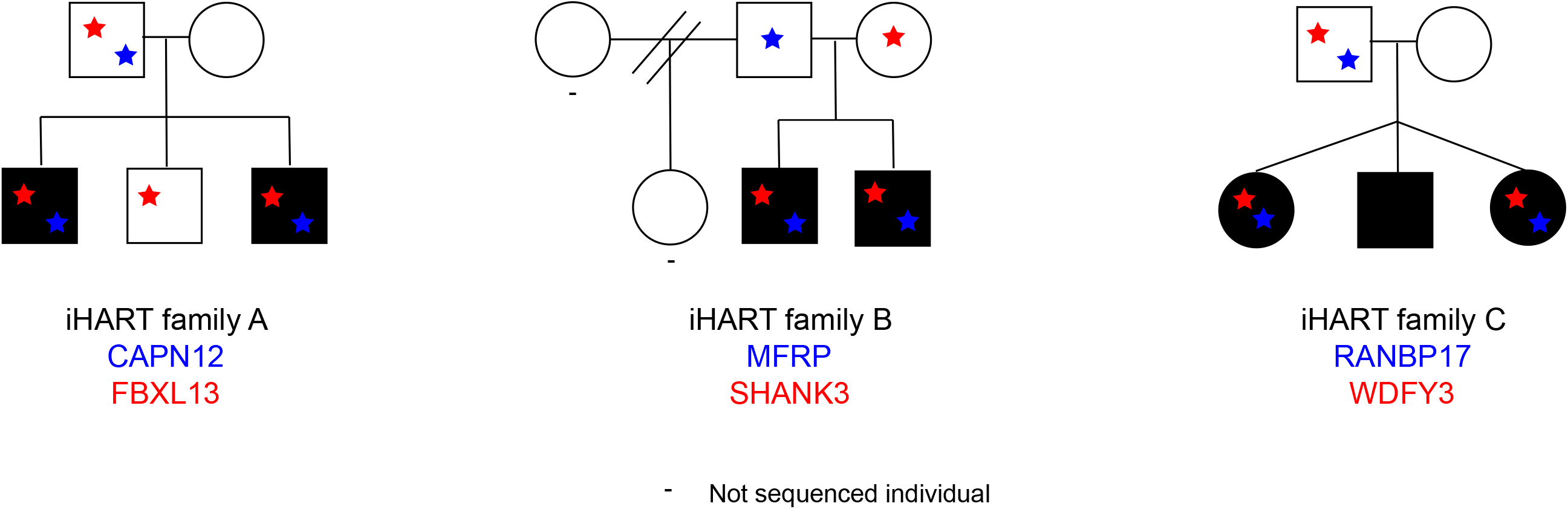
iHART family pedigrees where affected children inherited more than one protein truncating ASD risk gene.

Finally, we investigated the effect of rare inherited and rare *de novo* variants in these 152 known ASD risk genes on critical phenotypic measures describing affected children’s cognitive, social, and motor development. We observed significantly lower verbal IQ scores in all groups of affected children including carriers of rare inherited KARG variants (p=0.001), *de novo* carriers (p=0.02), and non-carriers (p<2×10^-16^, Figure 7A). Interestingly, we found significantly higher than population average non-verbal IQ scores (Raven population average = 100) in the affected non-carriers (p=1×10^-7^, Figure 7B), consistent with IQ lowering effects observed with rare variants in other studies (Satterstrom et al. 2020; Weiner et al. 2017). Both affected non-carriers and rare inherited carriers showed significantly delayed first steps (affected non-carriers p=1×10^-9^, rare inherited carrier p=0.03, Figures 7C), first words (affected non-carriers p<2×10^-16^, rare inherited carrier p<2×10^-16^, Figures 7D), and first phrases (affected non-carriers p<2×10^-16^, rare inherited carrier p<2×10^-16^, Figures 7E). We observed the largest effect of rare inherited KARG variants on language development (Figures 7D and 7E), specifically on the “first phrase” milestone achievement, with a median delay of 30 months compared to unaffected children from the same cohort (Figure 7E). We observed a similar significant trend for rare *de novo* carriers, with a median delay of 22 months (Figure 7E). As expected, all groups of affected children showed significantly higher SRS total raw scores compared to unaffected children from the same cohort (Figure 7F). We did not find significant differences among the three affected groups for any of the tested measures (Kruskal-Wallis test, Methods).

**Figure 7.**
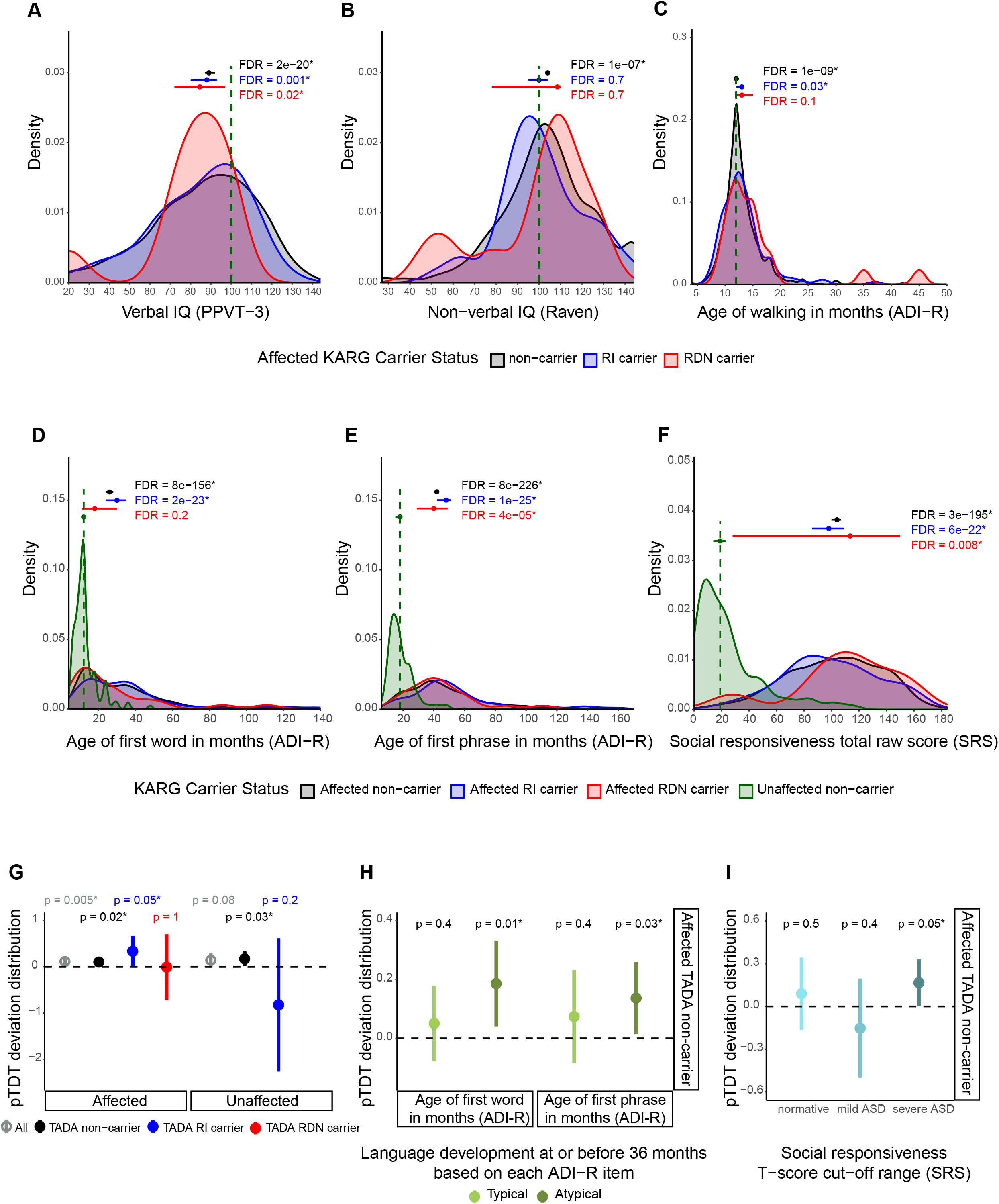
Phenotypic effect of rare and common risk for ASD in iHART affected children. (A-F) Phenotypic distributions in iHART affected children, stratified by those carrying rare inherited (RI) or rare *de novo* (RDN) variants in known ASD risk genes (KARG) and those not carrying such variants. (A) Verbal IQ distribution for affected children. Standard scores from PPVT-3 (Peabody Picture Vocabulary Test, 3rd edition) were used and compared to a standard hypothesized median value of 100. Affected non-carrier: n = 767, mu = 89; affected RI carrier: n = 70, mu = 88; affected RDN carrier: n = 10, mu = 84.5. (B) Non-verbal IQ distribution for affected children. Standard scores from the Raven test were used and compared to a standard hypothesized median value of 100. Affected non-carrier: n = 987, mu = 104; affected RI carrier: n = 98, mu = 100; affected RDN carrier: n = 12, mu = 108.5. (C) Age of walking distribution for affected children whose ADI-R (Autism Diagnostic Interview-Revised) data was available. A standard hypothesized median value of 12 months, observed in the normative cohort of the WHO Multicentre Growth Reference Study (MGRS) (Onis 2006), was used for comparison. Affected non-carrier: n = 1180, mu = 12; affected RI carrier: n = 120, mu = 13; affected RDN carrier: n = 21, mu = 13. (D) Age of first word for affected children whose ADI-R data was available. Affected non-carrier: n = 1223, mu = 26; affected RI carrier: n = 126, mu = 30; affected RDN carrier: n = 21, mu = 18; unaffected non-carrier: n = 295; mu = 12. (E) Age of first phrase for affected children whose ADI-R data was available. Affected non-carrier: n = 1220, mu = 42; affected RI carrier: n = 126, mu = 48; affected RDN carrier: n = 21, mu = 40; unaffected non-carrier: n = 283; mu = 18. (F) SRS (Social Responsiveness Scale) total raw score distribution for affected and unaffected iHART children. Affected non-carrier: n = 709, mu = 104; affected RI carrier: n = 73, mu = 98; affected RDN carrier: n = 8, mu = 113.5; unaffected non-carrier: n = 153; mu = 19. Non-carrier distributions are depicted in black, RI carrier ones in blue, and RDN ones in red. Standard hypothesized median values used for each phenotypic comparison are reported as green vertical lines. For panels D to F, these median values were derived from the corresponding phenotypic distributions for iHART unaffected non-carriers (shown in green) for whom phenotypic data was available. Each group of affected children was tested for difference among its phenotypic median value and the standard hypothesized one by two-sided one-sample sign test. The three tests performed for each phenotypic measure were corrected by multiple hypothesis testing using the Benjamini and Hochberg (BH) procedure as FDR (false discovery rate)-controlling method. An asterisk is used to denote an FDR < 0.05. Median values together with their 95% confidence intervals are represented by horizontal point ranges over the distributions. (G-I) ASD polygenic risk transmission stratified by ASD risk gene carrier type and association with language and social impairment. (G) pTDT in affected and unaffected children, tested for all subjects (All - affected: n = 1231; unaffected: n = 288) and stratified by those carrying rare inherited (TADA RI carrier - affected: n = 70; unaffected: n = 8) or rare *de novo* (TADA RDN carrier - affected: n = 12) variants in the TADA ASD risk genes and those without such variants (non-carriers - affected: n = 1149; unaffected: n = 279). (H) pTDT in affected non-carriers of variants in the TADA ASD risk genes stratified by those with atypical (age of first word: n = 407; phrase: n = 564) language development (i.e., whose ages of first word and phrase indicate that developmental concerns are present at or before 36 months, according to corresponding ADI-R, Autism Diagnostic Interview-Revised, scoring system for the two items) and those with typical (age of first word: n = 432; phrase: n = 275) language development. (I) pTDT in affected non-carriers of variants in the TADA ASD risk genes stratified by the degree of social impairment based on SRS (Social Responsiveness Scale) T-score cut-off ranges. T-scores (based on child’s sex) equal to 59 or less were considered as normative (n = 122), those 60 to 75 were considered mild ASD (n = 70), and those equal to or greater than 76 were considered severe ASD (n = 275) (Lyall et al. 2014). Polygenic transmission disequilibrium is shown on the y axis of each panel as point ranges of the standard deviation on the mid-parent distribution mean values together with their 95% confidence intervals. The probability of each pTDT deviation distribution mean being equal to zero was tested by two-sided one-sample t-test. An asterisk is used to denote a p-value < 0.05.

Overall, these finding show that contributory rare protein truncating variants from known ASD risk genes in affected children are inherited from unaffected parents. Furthermore, some affected children likely have an oligogenic burden. Lastly, ASD phenotypes, particularly non-verbal IQ and language development, varies by the form of known ASD risk gene inheritance.

### Polygenic Risk is Over Transmitted in Affected Children with Inherited Variants and Associated with ASD Phenotype

Given the pleiotropy associated with *de novo* variants and complex patterns of inheritance including oligogenic inheritance of rare PTVs, we next assessed the degree to which polygenic risk (PGR) contributes to ASD susceptibility in those harboring either *de novo* or inherited rare variants in the 74 TADA genes identified in this cohort. This is especially important, given the observation of a contribution from rare inherited variants from unaffected parents. We used the polygenic transmission disequilibrium test (pTDT) (Weiner et al. 2017), an extension of the TDT (Spielman, McGinnis, and Ewens 1993) to polygenic risk scores, which compares the mean of the offspring polygenic risk score (PRS) distribution to its parentally derived expected value. We confirmed overall, that affected children have an over transmission of ASD polygenic risk (two-sided, one-sample t-test, p=0.005, Figure 7G, Methods) compared with unaffected siblings, to whom no over-transmission of PRS is observed (p=0.08, Figure 7G).

To compare the contributions of PRS in the different variant classes, we stratified both affected and unaffected children into those with rare *de novo* PTV and missense variants in ASD risk genes and those with rare inherited PTVs (Figure 7G). Among affected children with an inherited protein truncating variant from statistically supported ASD risk genes identified in this study, we observed a significant over-transmission of PGR (p=0.05). This additive effect of ASD PGR to ASD rare inherited variation was not observed for PRS for schizophrenia (SCZ), bipolar disorder (BD), and educational attainment (EA) (Figure S6A, Methods). In contrast, affected children harboring a rare *de novo* PTV and/or missense variant (p=1, Figure 7G) and unaffected children with an inherited PTV variant do not exhibit an over-transmission of PRS (p=0.2, Figure 7G). We observed an over-transmission of ASD common risk in both affected (p=0.02) and unaffected (p=0.03) non-carriers (Figure 7G).

To investigate the effect of common risk for ASD on phenotypes related to cognitive and social impairment, and motor and language development, we performed pTDT analysis for affected non-carriers divided into typical and atypical groups based on critical cutoffs for each measure of interest (Methods). Interestingly, we observed the significant over-transmission of ASD PRS in children showing an atypical language development (age of first word and phrase greater than 24 and 33 months, respectively, suggesting developmental concerns before or at 36 months) (first word p=0.01, first phrase p=0.03, Methods, Figure 7H) and not in those with a typical language development (age of first word and phrase before or at 24 and 33 months, respectively, Methods) (first word p=0.4, first phrase p=0.4, Methods, Figure 7H). We did not find the same effect when testing PRS for SCZ, BD, and EA (Figure S6B, Methods). These findings suggest that the influence of common risk on child language development is specific to ASD polygenic risk.

In addition, we stratified the affected non-carriers by their social behavior and social interaction skills using T-score cut-off ranges from the Social Responsiveness Scale (SRS). As expected, the social impairment of these affected children falls within a spectrum: despite their diagnosis. Some children show neurotypical social skills; in contrast, others show deficiencies in reciprocal social behavior that would be categorized as mild or severe ASD (Figure 7I, Methods). Interestingly, we found a significant over-transmission of ASD PRS only in children with severe social impairment (p=0.05, Figure 7I, Methods) and not in those with mild social impairment (p=0.4, Figure 7I). We did not observe any difference in the contribution of common risk to social behavior among these groups when testing PRS for SCZ, BD, and EA (Figure S6C, Methods). These results confirm social behavior impairment as a core diagnostic domain for ASD and suggest its association with ASD PRS. Finally, we did not note any suggestive link between ASD PGR over-transmission and cognitive impairment (Figures S6D and S6E, Methods) or motor development (Figure 5E, Methods).

Taken together, these findings indicate that polygenic ASD risk is over transmitted to affected children, especially those with inherited protein truncating variants in ASD risk genes, and far less so to those harboring rare *de novo* variants. Furthermore, over transmission of common ASD risk is observed in ASD children with more severe language and social impairment, connecting language impairment to common genetic risk for ASD. This extends our understanding of ASD risk transmission to demonstrate co-inheritance of both rare and common risk in ASD probands.

## Discussion

Most genetic studies on ASD focus on simplex families, in which only one child is affected and optimally isolates strongly acting, rare *de novo* variants (O’Roak et al. 2012; Iossifov et al. 2014; O’Roak et al. 2014; De Rubeis et al. 2014; Krumm et al. 2015; Sanders et al. 2015; Turner et al. 2016; Stessman et al. 2017; Satterstrom et al. 2020). These large sequencing efforts on simplex cohorts have revolutionized ASD research, elucidating ASD genetic architecture and facilitating the identification of high-confidence risk genes for the disorder. However, our previous study on the first release of the iHART cohort, which was the largest cohort composed of multiplex families with more than one affected child, detected for the first time a strong additional risk contribution from rare inherited variants which was not detected in simplex families (Ruzzo et al. 2019). This previous study suggested a different genetic architecture for multiplex families which subsequently led to the discovery of new ASD risk genes whose association signal was derived from rare inherited variation and that impacted distinct biological processes from those hit by rare *de novo* variation (Ruzzo et al. 2019). WGS analysis on the extended iHART/AGRE cohort, which is two times the size, provided the opportunity to (1) validate and extend the list of ASD risk genes supported by rare inherited variation, (2) confirm the distinct ASD genetic architecture of multiplex families in a sufficiently large cohort, and (3) thoroughly assess the additive effects of both common and rare variation for ASD and its phenotypic spectrum.

We identified 7 novel ASD risk genes, 5 of which are impacted by a significantly higher proportion of rare inherited PTVs versus rare *de novo* variants. Our extended cohort analysis also validated two recently identified ASD risk genes, KMT2A and MED13L, and confirm that their association signal is mainly supported by rare *de novo* variation (Satterstrom et al. 2020). The overall yield of new risk genes defined by the presence of rare variants in this study likely reflects diminishing returns of novel ASD risk genes with increasing sample sizes, as has been projected for exome sequencing studies (Coe et al. 2019). We also did not identify an increased number of non-coding rare inherited, private inherited, or rare *de novo* variants in affected children, despite focusing on putatively functional elements of the non-coding genome such as brain-specific co-localized regions and TFBSs (Xu et al. 2020; Gheorghe et al. 2019; Vorontsov et al. 2018). Non-coding ASD genetic risk in simplex families has been identified from large DNase I hypersensitivity regions (Turner et al. 2016), TFBSs within the distal promoters of conserved loci (An et al. 2018), and non-coding variants predicted to be deleterious from deep-learning (Zhou et al. 2019). Our analysis agrees with previous analyses suggesting that sample sizes of roughly 8000 fully-phaseable children would be necessary to be able identify non-coding ASD genetic risk with 80% power (Werling et al. 2018).

Of the 74 genes associated with ASD (FDR < 0.1) from the current study, 39 converge in a significant indirect protein-protein interaction network. These significant seed genes include known ASD risk genes (*CHD8*, *GRIN2B*, *SHANK3*) (Ruzzo et al. 2019; Satterstrom et al. 2020) and newly identified risk genes (*FBXL13*, *SNCAIP*). Common genetic variation in FBXL13 was associated with sleep-wake cycle preferences (Veatch et al. 2017; Hu et al. 2016, 89; Lane et al. 2016), which is disrupted in ASD. *SCNAIP*, also known as Synphilin-1, has been extensively studied by promoting neurodegeneration in Parkinson’s disease when interacting with alpha-synuclein (Chung et al. 2001; Engelender et al. 1999, 1; Krüger 2004; Dashtipour et al. 2017). Parkinsonism and movement disorders including repetitive behaviors are recognized features of ASD (Qiu et al. 2010; Estes et al. 2011; Abbott et al. 2018; Starkstein et al. 2015; Bell, Wittkowski, and Hare 2019).

The predominant contribution of rare inherited variants to ASD risk in multiplex families is confirmed by our focused and comprehensive analysis on known ASD risk genes that had been previously identified in cohorts with predominantly simplex families (Satterstrom et al. 2020; Turner et al. 2017). Interestingly, affected children in multiplex families have a significant excess of rare inherited, but not rare *de novo* protein truncating variants in known ASD risk genes. We found that, in addition to ASD resulting from rare inherited variation, inherited multiplex ASD risk may in some cases involve transmission of more than one rare PTV to affected children. Affected children are more likely to carry one or two rare inherited PTVs in known ASD risk genes compared to unaffected children. A similar oligogenic burden in affected children was identified for *de novo* variants in simplex families (likely gene disrupting, severe missense variants, in fetal brain transcription factor binding sites, 3’ UTR and deletions that disrupt an exon) (Turner et al. 2017). We identified three pairs of siblings with two rare inherited PTVs in known ASD risk genes. In one family with two affected females and one affected male, the females had two inherited PTVs in known ASD risk genes (*WDFY3*, *RANBP17*) while the male, who had milder ASD symptoms, did not share these same variants. *RANBP17*, which was mutated in affected females of this family, showed a female-biased expression in human fetal brain (O’Brien et al. 2019), but not in human adult brain (Oliva et al. 2020). This specific case and sex specific expression of *RANBP17* supports the multiple threshold theory in females with ASD (Maenner et al. 2020; Fombonne 2009; Satterstrom et al. 2020; Werling and Geschwind 2013), which posits females require a greater burden of genetic risk to develop ASD compared to males.

Previous analyses investigating additivity between common polygenic and rare variation in simplex families showed that ASD common polygenic risk is over transmitted in affected children carrying a strongly acting *de novo* variant (PTVs and deletions) (Weiner et al. 2017). In this cohort, we showed that PRS is significantly over-transmitted to affected children harboring inherited protein coding rare variants in ASD risk genes. Because we observed a small, but significant over-transmission of ASD common risk in both affected and unaffected children who did not carry rare inherited ASD risk genes, our findings suggest the over-transmission of PRS in ASD acts in combination with rare inherited ASD genetic risk towards the development of ASD. Thus, PRS combines with rare variants to influence risk for ASD (Takahashi et al. 2020; Serdarevic et al. 2020) and may partially explain why parents with protein coding rare variants in ASD risk genes are asymptomatic, as they lack the polygenic ASD risk which was transmitted to their affected children.

Another important observation is that ASD PRS is not only over-transmitted to affected children with severe social impairment, but also those with atypical language development at or before 36 months. In contrast, PRS for SCZ, BD, and EA did not show a similar effect on language development and social behavior. Other recent work (in preprint form) found a positive association between ASD polygenic risk and earlier age of first word in affected children from SSC and SPARK (Warrier et al. 2021). However, this signal was no longer significant after accounting for presence of high-impact *de novo* variants and full-scale IQ (Warrier et al. 2021). Interestingly, results from another recent preprint, relevant to our SRS finding, show a positive association between polygenic risk for ASD and severity of social communication deficits, but not of social behavior impairment measured by SRS (Antaki et al. 2021). We did not observe any suggestive effect of ASD common risk on cognitive impairment or motor development, consistent with observations from other cohorts highlighting the importance of rare variation vis a vis the manifestation of these phenotypes (Weiner et al. 2017; Antaki et al. 2021).

The novel link between language delay and over-inheritance of PRS for ASD shown here supports the relevance of assessing accompanying language impairment in the context of ASD diagnosis. Moreover, these results suggest that indeed language impairment is a core feature of ASD and should not be a secondary consideration with regards to diagnosis, as it is in the current version of DSM-V. Since this is the first time an association between ASD PRS and language delay has been observed, it will be important to replicate it in additional cohorts. Nevertheless, this work highlights the value of multiplex family cohorts in ASD research where the effects of rare and common inherited variation are more easily observed and can be associated with neurobiologically relevant phenotypes, such as language or motor function.

## Data Availability

The Hartwell Foundation's Autism Research and Technology Initiative (iHART) will provide the whole genome sequencing data generated during this study upon request and approval of the data use agreement available at http://www.ihart.org.

## Acknowledgments

We thank the subjects and families from the Autism Genetic Resource Exchange (AGRE) who participated in this study, in addition to the Hartwell Foundation, and New York Genome Center. We thank Stephanie N. Kravitz, Cheyenne L. Schloffman, David Keller, Min Sun, Tor Solli-Nowlan, Sasha Sharma, Marlena Duda, Greg Madden McInnes, Ravina Jain, Valentı’Moncunill, Josep M. Mercader, Montserrat Puiggrò s, Anika Gupta, and David Torrents for technical support. We are grateful to all of the families at the participating SSC sites as well as the principal investigators (A. Beaudet, R. Bernier, J. Constantino, E. Cook, E. Fombonne, D. Geschwind, R. Goin-Kochel, E. Hanson, D. Grice, A. Klin, D. Ledbetter, C.Lord, C. Martin, D. Martin, R. Maxim, J. Miles, O. Ousley, K. Pelphrey, B. Peterson, J. Piggot, C. Saulnier, M. State, W. Stone, J. Sutcliffe, C. Walsh, Z. Warren, and E. Wijsman).

## Funding/Support

This work was supported by grants from The Hartwell Foundation and the NIH (R01MH100027, R01MH064547, S10OD011939, P50HD055784, UM1HG008901, P50DC018006, K08AG065519 R01MH064547, S10OD011939) and from the Stanford Precision Health and Integrated Diagnostics Center and from the Stanford Bio-X Center. We are grateful to The Hartwell Foundation for supporting the genetic sequencing and analyses of iHART. We thank the Hartwell Foundation for the opportunity to contribute to their mission of benefitting children, and for the financial support that enabled the whole-genome sequencing and analyses. We are grateful to the Simons Foundation for additional support for genome sequencing. We thank the New York Genome Center for conducting sequencing and initial quality control. We thank Amazon Web Services for their grant support for the computational infrastructure and storage for the iHART database. The Autism Genetic Resource Exchange (AGRE) is a program of Autism Speaks and was supported by grant NIMH U24 MH081810. The Simons Simplex Cohort data described in this study can be obtained through application to SFARI Base (https://base.sfari.org).

## Author Contributions

T.S.C., M.C., and S.A.A. contributed to the analytical plans, performed analyses, and interpreted results. J.K.L. selected and submitted samples for sequencing. J.Y.J. performed joint genotyping, VCF annotation, and data transfers. M.C., S.A.A., E.K.R., J.Y.J., L.K.W., and J.K.L. performed quality control checks. L.K.W. wrote scripts for data processing and helped interpret results. L.P.C., D.K.H., J.Y.J., E.K.R., and D.P.W. developed ARC. E.K.R. and L.P.C. contributed to analytical plans and performed TADA. A.G. and L.B. performed TADA follow up functional analyses. M.C. and J.K.L analyzed phenotypes. D.P.W. identified and supplied funding. T.S.C., M.C., and S.A.A. took the lead in writing the manuscript. All authors reviewed, edited, and approved the manuscript. D.H.G. and D.P.W. supervised the experimental design and analysis and interpreted results.

## Declaration of Interests

The authors declare no competing interests

## Supplemental Figure Legends

**Figure S1.**
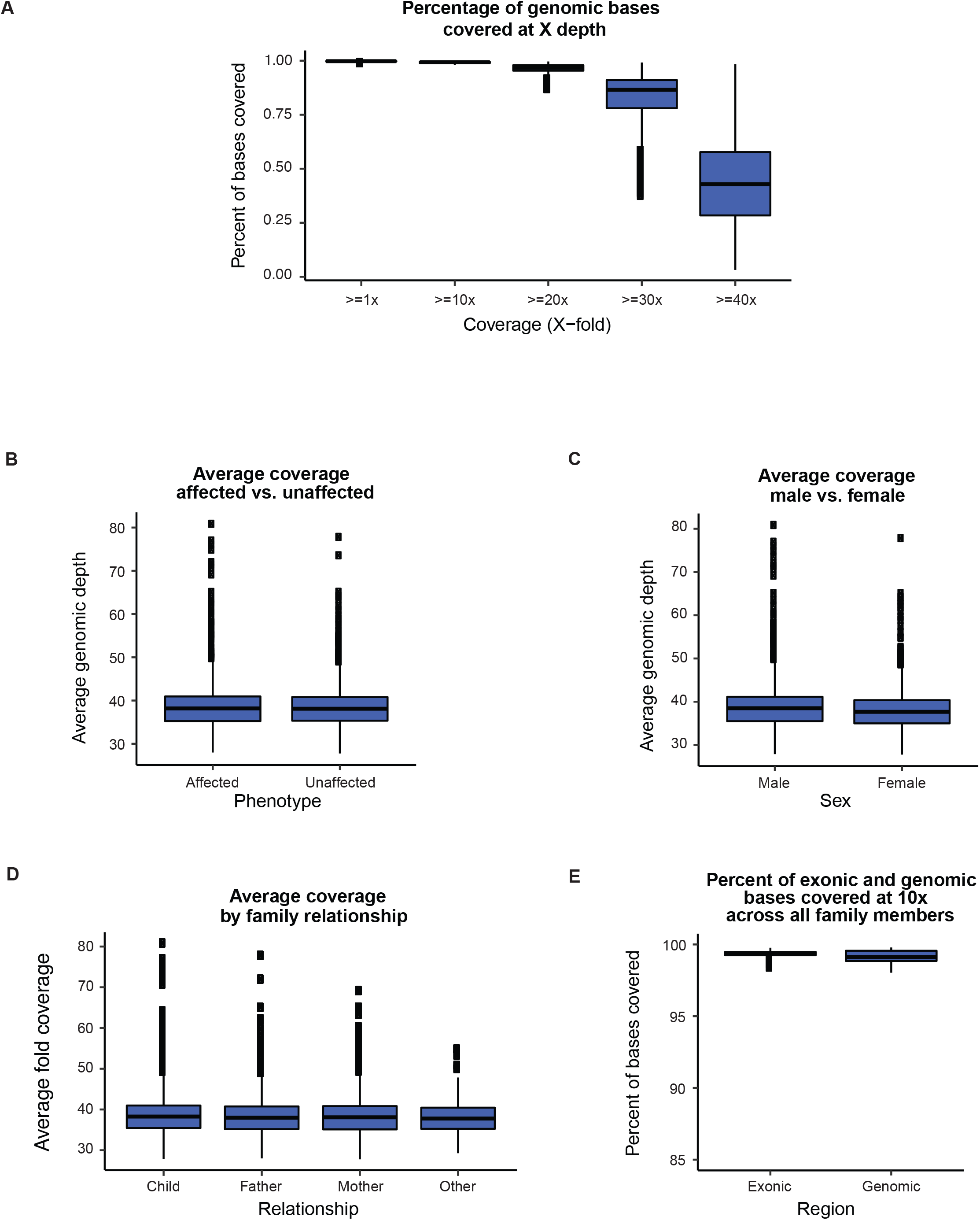
WGS coverage statistics for 4,551 iHART/AGRE samples. (A) For the 4,551 iHART samples with WGS data, the percent of genomic bases covered at 1X, 10X, 20X, 30X, and 40X bases. On average, 99.23 ± 0.35% of bases were covered at a depth of ≥ 10X. The average fold coverage per sample across the cohort and no differences in the categories of (B) ASD affectation status, (C) sex, or (D) family member type – where family member type was categorized as either Child (proband, sibling, MZ or DZ twin), Father, Mother, or Other (e.g., cousin). (E) The percent of exonic and genomic bases covered at ≥ 10x across all family members present in the 865 fully-phaseable iHART families. Exonic regions are those that are annotated as protein-coding exons (> 75MB) in Gencode V19. All non-N bases in the reference genome (>2.8Gb) were considered genomic regions.

**Figure S2.**
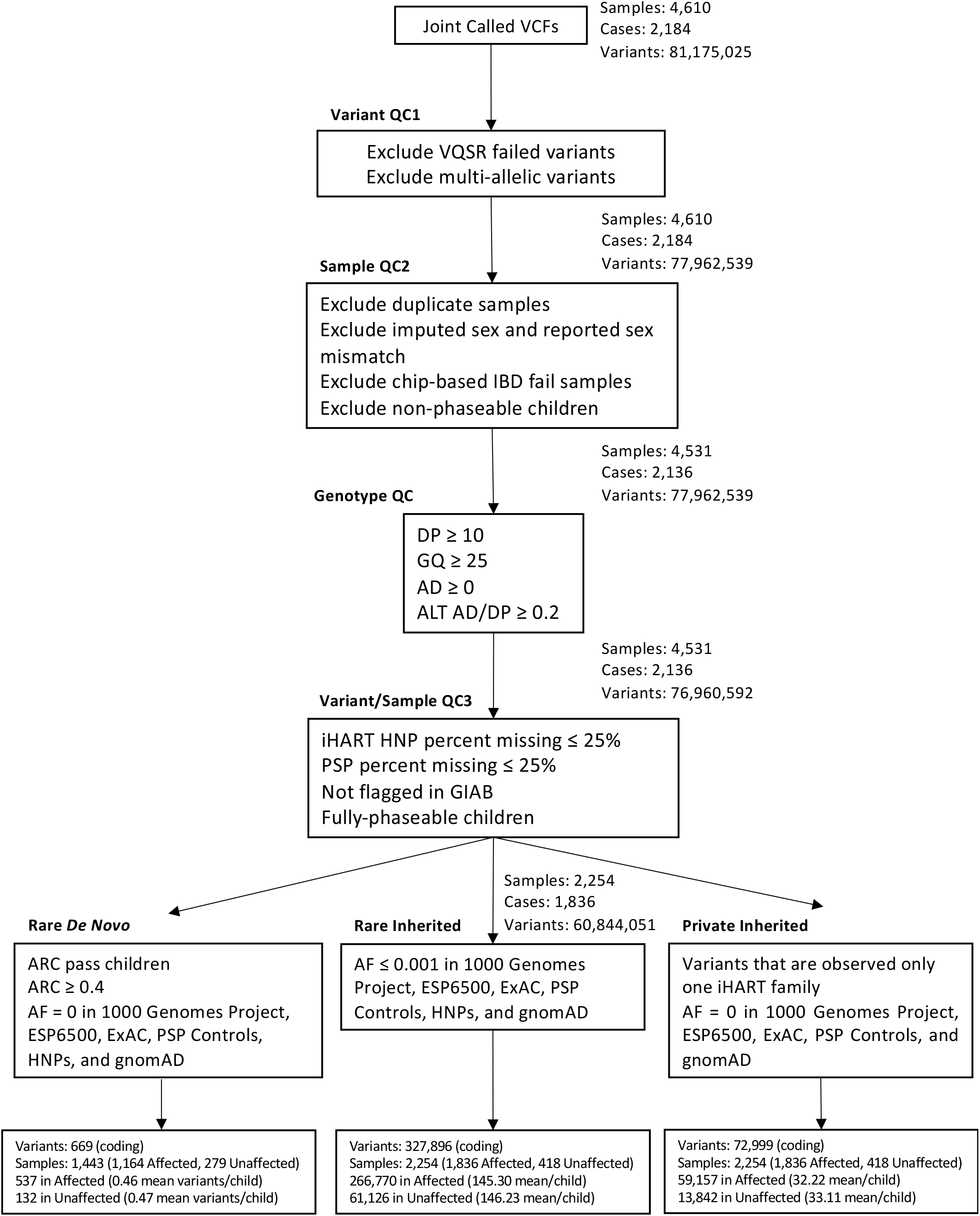
Whole genome sequencing pipeline overview. An overview of the variant, sample, and genotype quality control conducted on the raw joint called variants to obtain the rare *de novo*, rare inherited, and private inherited variants used in downstream analyses. Abbreviations: VCF (Variant Call Format), QC (Quality Control), VQSR (Variant Quality Score Recalibration), IBD (Identity by Descent), DP (Read Depth), GQ (Genotype quality), AD (Allele Depth), HNP (Healthy Non-phaseable), PSP (Progressive Supranuclear Palsy), GIAB (Genome in a Bottle Consortium).

**Figure S3.**
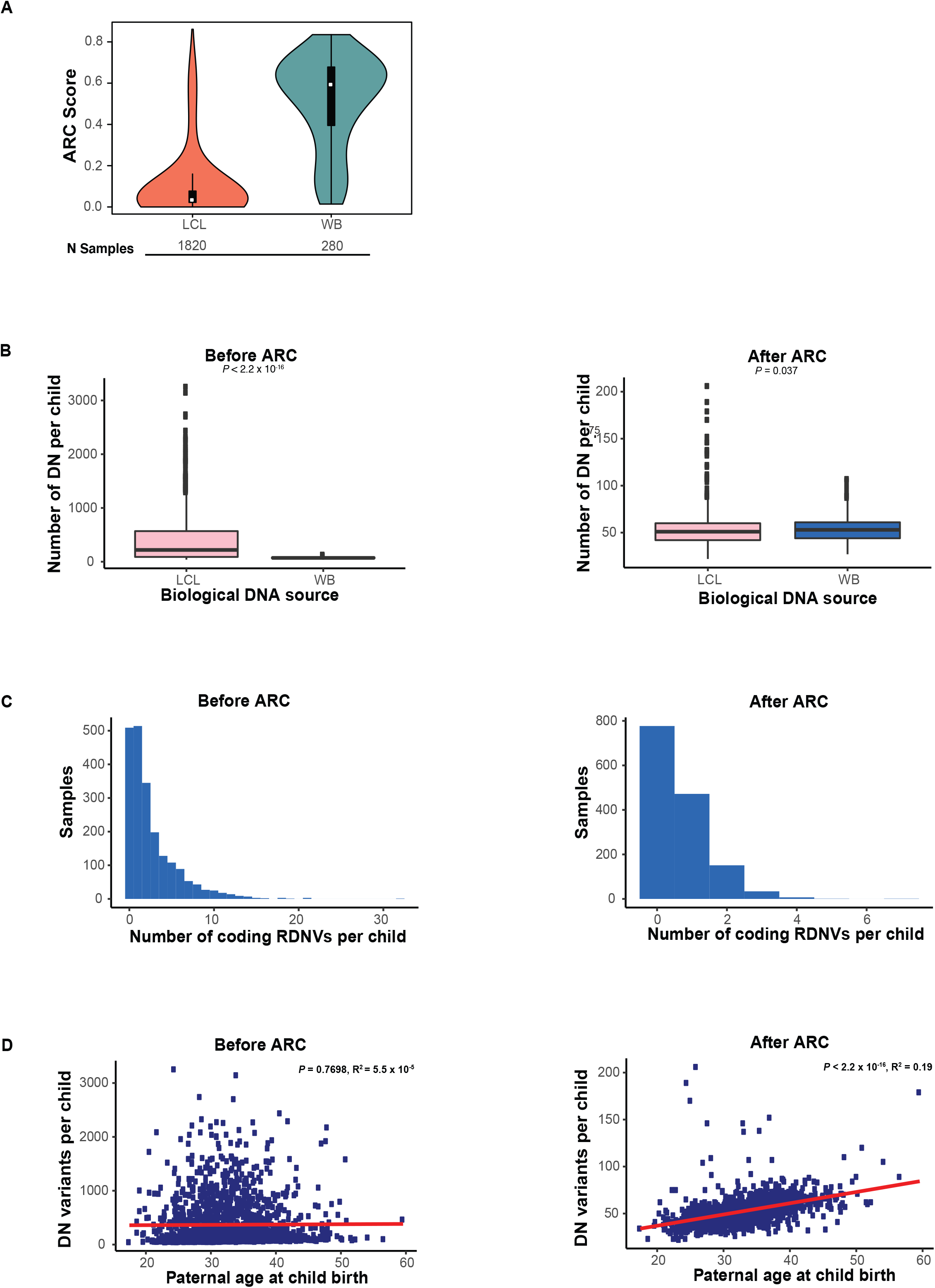
Rare *de novo* variants (RDNVs) in iHART samples before and after Artifact Removal by Classifier (ARC). (A) Distribution of raw *de novo* variant ARC scores from 2,100 fully-phaseable non-monozygotic (MZ) twin samples. Red represents samples derived from lymphoblastic cell lines (LCL) and green represents samples derived from whole blood (WB). (B) In LCL (pink) and WB (blue) fully-phaseable (non-MZ twin) samples, the number of rare *de novo* variants identified per sample before ARC (N = 2,100 samples) and the number of rare *de novo* variants identified in LCL (pink) and WB (blue) fully-phaseable (non-MZ twin) samples after ARC (variants with an ARC score < 0.4 are filtered out) and after excluding ARC outlier samples (samples with > 90% DNs removed by ARC) (N = 1,443). After ARC, there is a less significant difference in the rate of rare *de novo* variants based on the biological sequencing source (LCL mean = 52.6 and WB mean = 53.6; LCL median = 51 and WB median = 53). The Wilcoxon rank sum test was used to evaluate the difference in DN rates between the biological sequencing source (LCL versus WB). (C) Histograms displaying the number of rare *de novo* coding variants per fully-phaseable sample. Before ARC, coding RDNVs come from 2,100 fully-phaseable non-MZ twin samples; after ARC (which filters out variants with an ARC score < 0.4), variants are derived from samples excluding ARC outliers (samples with > 90% DNs removed by ARC) and MZ twins (n = 1,443). (D) The correlation between paternal age and the rate of rare *de novo* variants before and after ARC. This analysis considers 1,677 fully-phaseable ASD children (excluding MZ twins) for which paternal age was known before ARC and 1,157 fully-phaseable ASD children (excluding MZ twins and ARC outliers) for which paternal age was known after ARC. The linear regression line is in red. On the left, the graph shows the number of raw rare *de novo* variants (SNVs and indels) per child by paternal age (years) at the time of the participant’s birth. On the right, the graph shows the number of rare *de novo* variants (SNVs and indels) per child identified after running ARC by paternal age (years) at the time of the participant’s birth.

**Figure S4.**
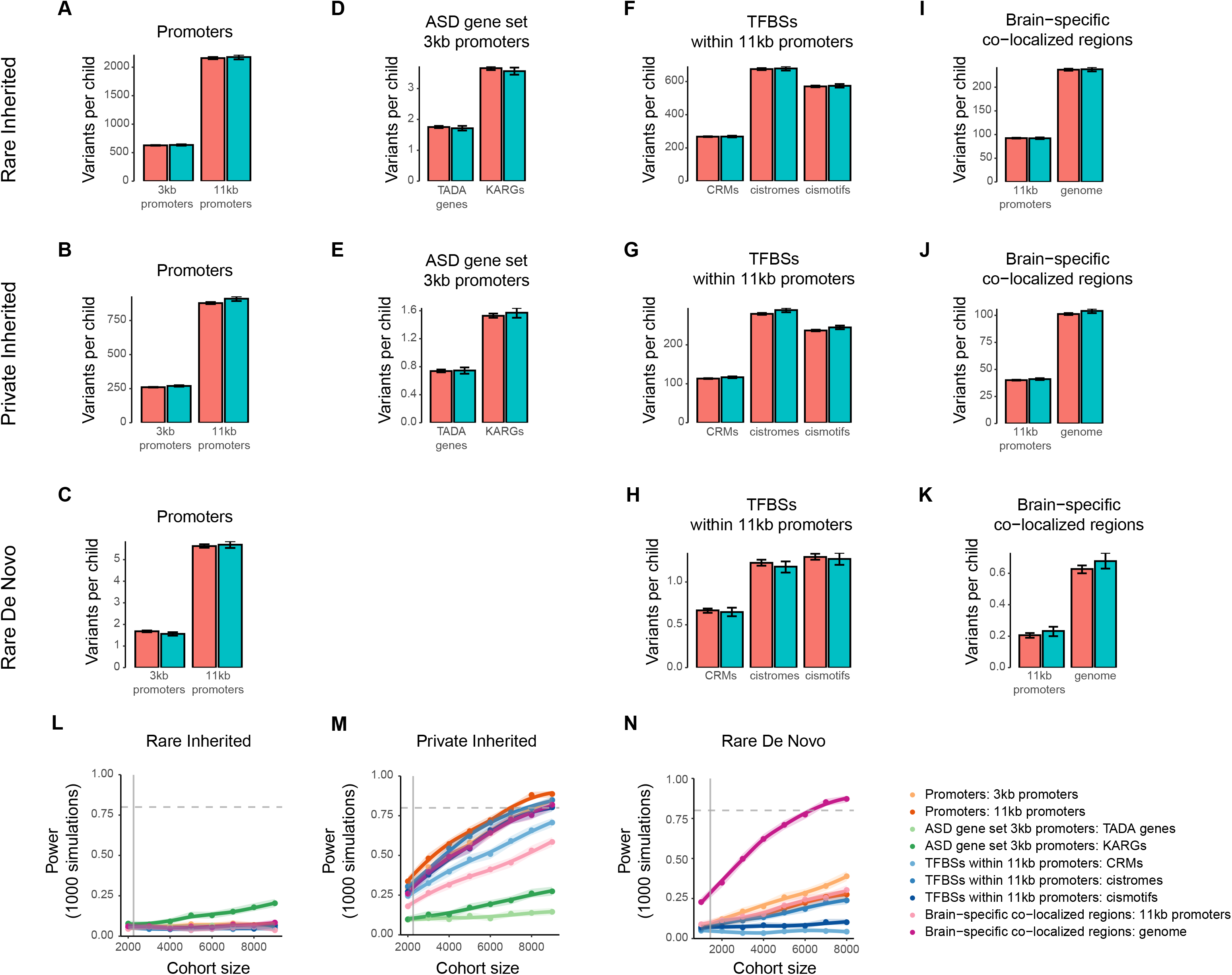
(A-K) Number of non-coding variants per fully-phaseable child in affected (salmon) and unaffected (turquoise) children. Mean ± standard error variant count per child are shown. (A-D) For promoter regions, rare inherited non-coding variants in 1,836 affected versus 418 unaffected children (A), private inherited non-coding variants in 1,836 affected versus 418 unaffected children (B), and rare *de novo* non-coding variants in 1,164 affected versus 279 unaffected children (C). (D-E) For promoter regions of ASD risk genes, rare inherited non-coding variants in affected versus unaffected children (D), and private inherited non-coding variants affected versus unaffected children (E). (F-H) For transcription factor binding sites in promoter regions, rare inherited non-coding variants in affected versus unaffected children (F), private inherited non-coding variants affected versus unaffected children (G), and rare *de novo* non-coding variants in affected versus unaffected children (H). (I-K) For brain-specific co-localized regions, rare inherited non-coding variants in affected versus unaffected children (I), private inherited non-coding variants affected versus unaffected children (J), and rare *de novo* non-coding variants in affected versus unaffected children (K). (L-N) Curves showing power at increasing sample sizes for the quantitative burden testing through logistic regression of rare inherited (L), private inherited (M), and rare *de novo* (N) non-coding variants. Each curve corresponds to a tested variant set (see color-coded legend), with individual points representing power values specific to the “variant count per child” predictor, computed based on 1,000 simulations, for the observed ASD odds ratio (OR) and at specific sample sizes. 95% confidence intervals for these power values are depicted as ribbons. The dashed grey horizontal line marks the 0.8 standard threshold used for power while the solid grey vertical line shows the current sample size for the specific variant set (2,254 fully-phaseable children for rare and private inherited non-coding variants and 1,443 children for rare *de novo* non-coding variants).

**Figure S5.**
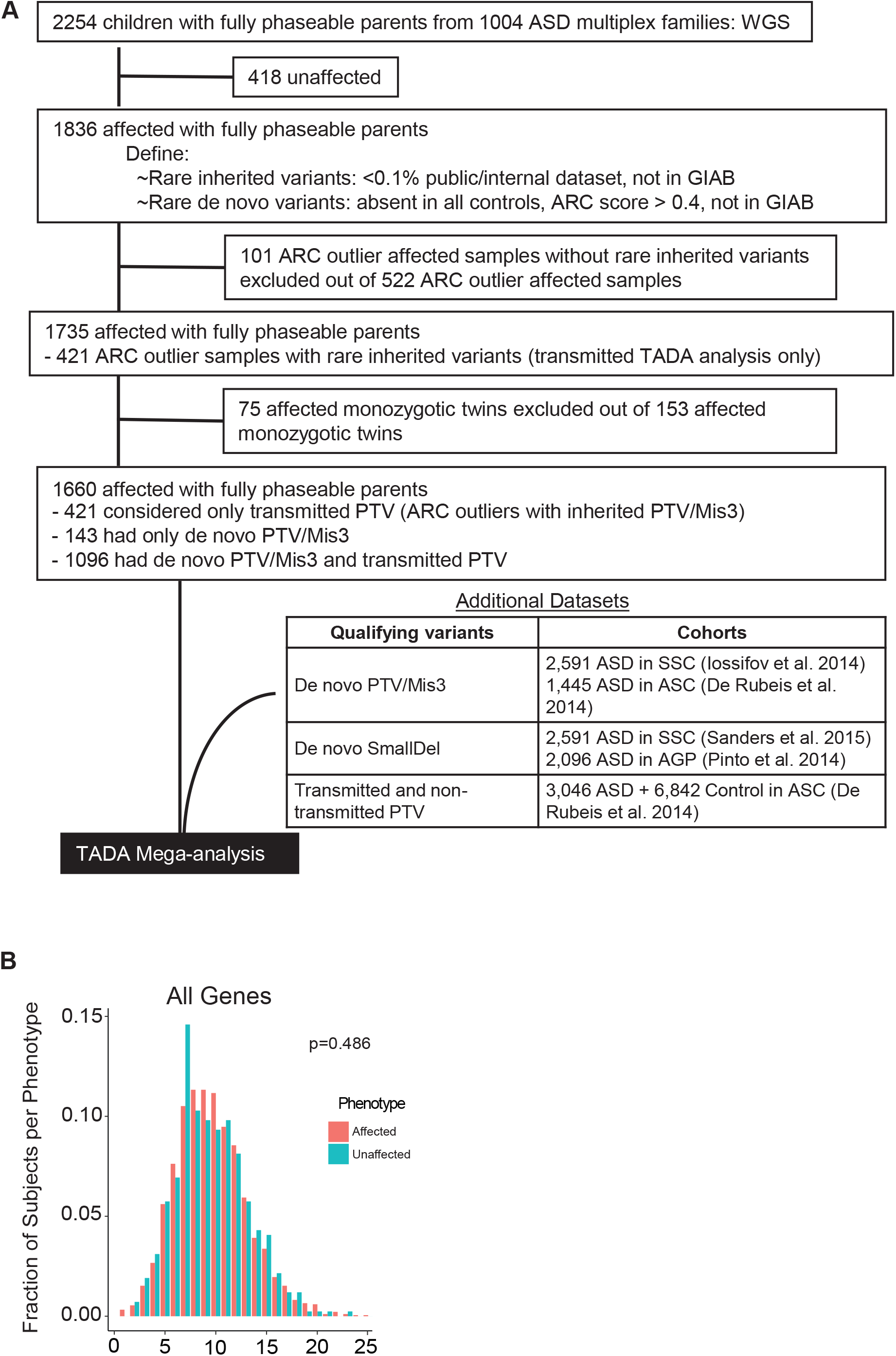
(A) Detailed overview of iHART subjects and additional datasets included in the TADA analysis. Initial dataset included 2,254 children with fully-phaseable parents from 1,004 ASD multiplex families. Subjects were removed if they were unaffected, were considered ARC outliers or were a monozygotic twin. Subsets of the 1660 affected children were included in TADA for *de novo* or inherited analysis. Additional datasets used for TADA mega-analysis are shown. (B) Fraction of subjects and number of genes with protein truncating variants from all genes.

**Figure S6.**
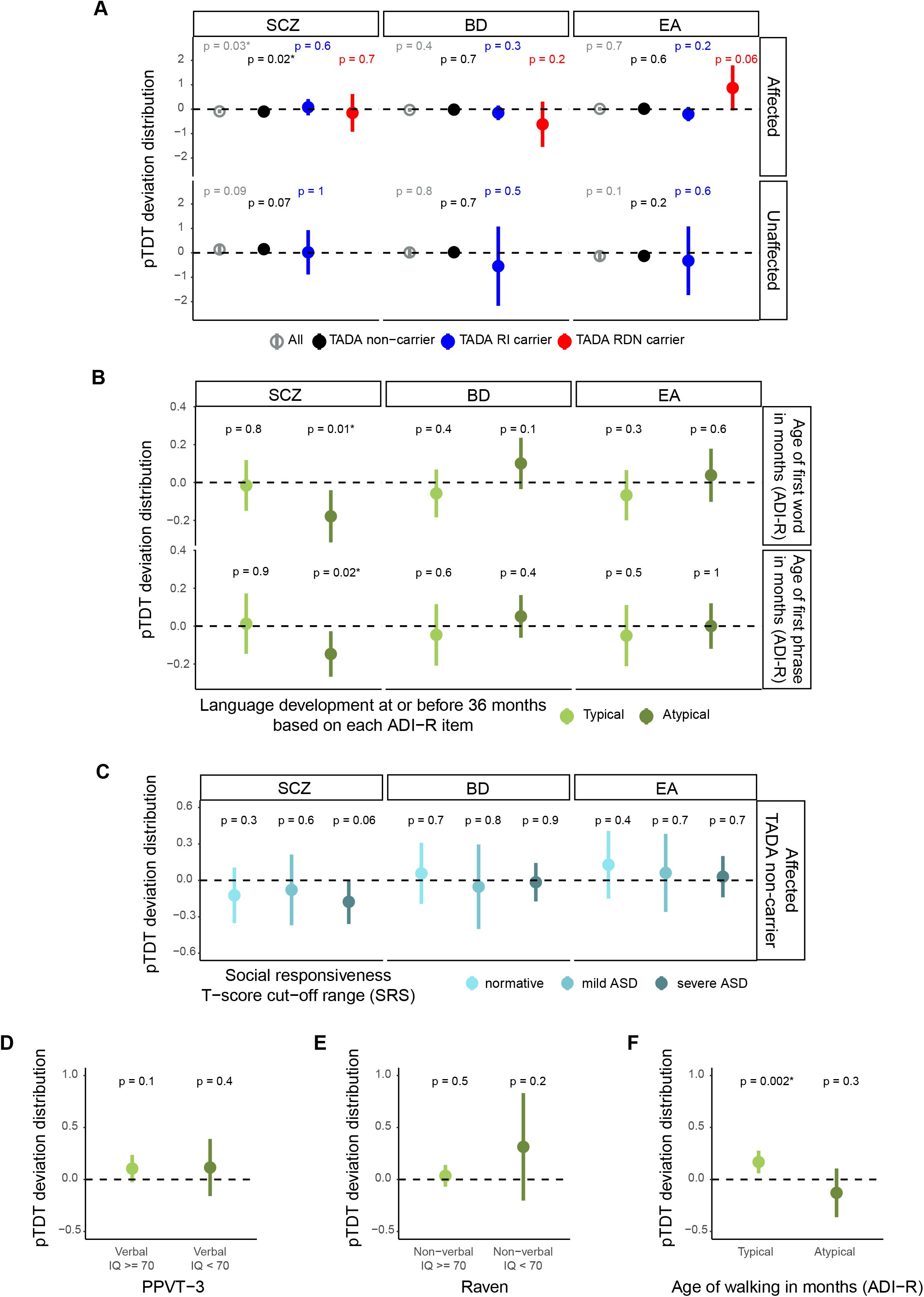
Polygenic risk for schizophrenia (SCZ), bipolar disorder (BD), and educational attainment (EA) in iHART and ASD pTDT stratified by cognitive and motor impairment. (A) pTDT for SCZ, BD, and EA in affected and unaffected children, tested all together (All - affected: n = 1231; unaffected: n = 288) and stratified by those carrying rare inherited (TADA RI carrier - affected: n = 70; unaffected: n = 8) or rare *de novo* (TADA RDN carrier - affected: n = 12) variants in the TADA ASD risk genes and those without such variants (non-carriers - affected: n = 1149; unaffected: n = 279). (B) pTDT for SCZ, BD, and EA in affected non-carriers of variants in the TADA ASD risk genes stratified by those with atypical (age of first word: n = 407; phrase: n = 564) language development (i.e., whose ages of first word and phrase indicate that developmental concerns are present at or before 36 months, according to corresponding ADI-R, Autism Diagnostic Interview-Revised, scoring system for the two items) and those with typical (age of first word: n = 432; phrase: n = 275) language development. (C) pTDT for SCZ, BD, and EA in affected non-carriers of variants in the TADA ASD risk genes stratified by the degree of social impairment based on SRS (Social Responsiveness Scale) T-score cut-off ranges. T-scores (based on child’s sex) equal to 59 or less were considered as normative (n = 122), those 60 to 75 were considered mild ASD (n = 70), and those equal to or greater than 76 were considered severe ASD (n = 275) (Lyall et al. 2014). (D-E) pTDT for ASD in affected non-carriers of variants in the TADA ASD risk genes stratified by those with (IQ < 70) or without (IQ >= 70) cognitive impairment, based on their verbal IQ scores from PPVT-3 (Peabody Picture Vocabulary Test, 3rd edition) (D) and non-verbal IQ ones from the Raven test (E). Verbal IQ >= 70: n = 430; Verbal IQ < 70: n = 106; Non-verbal IQ >= 70: n = 682; Non-verbal IQ < 70: n = 39. (F) pTDT for ASD in affected non-carriers of variants in the TADA ASD risk genes stratified by those with typical (age of walking in months < 16, n = 669) or atypical (age of walking in months >= 16, n = 135) motor development. The age of 16 months chosen for stratification is the 97th percentile estimate for milestone achievement age in the normative cohort of the WHO Multicentre Growth Reference Study (MGRS) (Onis 2006). Polygenic transmission disequilibrium is shown on the y axis of each panel as point ranges of the standard deviation on the mid-parent distribution mean values together with their 95% confidence intervals. The probability of each pTDT deviation distribution mean being equal to zero was tested by two-sided one-sample t-test. An asterisk is used to denote a p-value < 0.05. More details provided in the Methods section.

Table S1. Sample demographics, family relationships and whole genome sequencing metrics

Table S2. Results from non-coding variant quantitative burden testing through logistic regression. For each non-coding variant set tested, total number of variants, significance of the logistic regression model built, and significance and ASD odds ratio (OR) for the three model predictors (variant count per child, child sex, and sample source of DNA used for whole-genome sequencing) are shown. Male sex and lymphoblastoid cell line (LCL) sample source of DNA were used as reference values for the two categorical variables. ASD OR for the variant count per child predictor represents the change in ASD odds for one unit increase of variant count per child.

Table S3. Results from power analysis for non-coding variant quantitative burden testing through logistic regression. For each non-coding variant set tested, power values for quantitative burden testing through logistic regression and specific to the variant count per child predictor are reported. Power was computed with a posthoc approach (for the observed ASD OR and at the current sample size) using three different simulation strategies (simr powerSim() function, “sample” custom function, and “random” custom function). Power was estimated over 1,000 simulations. A description of the simulation strategies can be found in the related methods’ section. 95% confidence intervals for the power values are shown within parentheses.

Table S4. Detailed TADA results including qualifying variants, cohorts included, results, and results by variant class

Table S5. Comparison table of TADA ASD risk genes, Known ASD risk gene, and development disorder risk gene list. A “1” indicates the inclusion of a gene within the gene list, while a “0” represents the absence of a gene.

## Methods

### LEAD CONTACT AND MATERIALS AVAILABILITY

The Hartwell Foundation’s Autism Research and Technology Initiative (iHART) will provide the whole genome sequencing data generated during this study upon request and approval of the data use agreement available at http://www.ihart.org. Lead Contact, Daniel Geschwind can be contacted for any further information or requests for resources and reagents.

### EXPERIMENTAL MODEL AND SUBJECT DETAILS

#### ASD multiplex family samples

As a result of this study being limited to previously-existing coded data and specimens, the UCLA and Stanford IRBs considered it “Not human subjects research” and thus exempt from review. Subjects originated from the Autism Genetic Resource Exchange (AGRE) (Lajonchere, 2010) and were carefully selected from families including at least two members with ASD. Sequencing was not performed on patients with known genetic causes of ASD or syndromes overlapping with ASD-features. Table S1 contains a complete list of sequenced samples.

Of subjects selected from the Autism Genetic Resource Exchange (AGRE), 4,551 individuals from 1,004 ASD families passed quality control. Table S1 provides information about these 4,551 samples, including sex, ethnicity, phenotype, and familial relationship. Our variant analysis included 4,531 individuals (2,136 affected and 2,395 unaffected); 20 children for whom both biological parents were not sequenced were excluded. If not otherwise specified, our analyses included a subset of 2,254 children (1,836 affected and 418 unaffected) whose parents had both undergone sequencing.

The Rutgers University Cell and DNA Repository (RUCDR; Piscataway, NJ) provided our purified DNA. Whenever possible, whole blood DNA was used; for many samples, however, only lymphoblastoid cell line (LCL) DNA was extracted at the time of recruitment.

#### Control cohorts

We refer to several control cohorts throughout the manuscript that we have used to assess variant frequencies in non-ASD samples. These cohorts are described below.

##### Publicly available databases

Annotations from publicly available databases were obtained from ANNOVAR and include: the 1000 genomes project (1000g2015aug_all) (Auton et al., 2015), gnomAD (version 2.0.2) (Karczewski et al., 2019), the Exome Aggregation Consortium (exac03nonpsych) (Lek et al., 2016), the NHLBI Exome Sequencing Project (esp6500siv2_all) (http://evs.gs.washington.edu/EVS/), and allele frequency estimates from whole-genome sequenced unrelated, healthy subjects (http://www.completegenomics.com/public-data/69-genomes/, cg46).

##### UCLA internal controls

The “UCLA internal controls” referred to throughout this manuscript are comprised a 379 unrelated, whole-genome sequenced samples with *Progressive Supranuclear Palsy* (PSP). ASD and PSP have no known etiological overlap or comorbidity.

##### Healthy Non-Phaseable (HNP) samples

In this manuscript, “HNPs” refers to 1,890 healthy iHART samples for which no biological parents have been sequenced (non-phaseable). These samples likely harbor genetic variants associated with ASD, and so provide allele frequency estimates that are generally more permissive when filtering for inherited risk variants.

### METHOD DETAILS

#### Whole-genome sequencing and data processing

Whole-genome sequencing was conducted on DNA samples through New York Genome Center (NYCC). Following DNA sample quality and quantity assessment, genotyping was performed using Illumina Infinium Human Exome-12 v1.2 or Infinium Human Core Exome microarrays (San Diego, CA). The genome-wide genotyping data was subsequently used to confirm sample identity and conduct sex checks in in PLINK v1.07 (Purcell et al., 2007). VerifyIDintensity (VII) (Jun et al., 2012) was used to assess contamination; samples with two or more modes of contamination greater than 3% were excluded from sequencing. Samples that passed identity and quality checks were then sequenced at NYGC using the Illumina TruSeq Nano library kits and Illumina’s HiSeq X (San Diego, CA) (Ruzzo et al. 2019). iHART WGS data were processed through the same bioinformatics pipeline, adapted from GATK’s best practices (DePristo et al., 2011; Van der Auwera et al., 2013). The bioinformatics processing steps are outlined in our previous manuscript (Ruzzo et al. 2019), with the only difference being that GATK tools were updated to version 3.4 and Picard MarkDuplicates tool to version 1.83.

#### Quality control assessment

WGS data underwent standard quality control checks to ensure the accuracy of sequencing and variants, as well as sample identity. Checks were conducted on samples for relatedness, duplicates, contamination, sequencing coverage, variant quality (using GATK’s VariantEval module, data not shown), concordance between genotyping chip and WGS data, and concordance between self-declared sex and observed biological sex. A total of 4,551 individuals from 1,004 ASD families, from the Autism Genetic Resource Exchange (AGRE) passed quality control (Table S1).

##### Whole-genome sequence coverage

We calculated genome-wide per-base sequencing coverage for each sample using SAMtools v1.2 (Li et al., 2009). Custom scripts were used to calculate (i) average coverage and (ii) percentage of the genome (excluding gap regions, downloaded from the UCSC table browser) covered at 1X, 10X, 20X, 30X, and 40X. On average across all samples, 99.2% of bases were covered at a depth of ≥10X (Figure S1A-E).

#### Variant inheritance classifications

Variants were classified into one of eight inheritance types: (i) *de novo*, (ii) paternally inherited, (iii) maternally inherited, (iv) missing, (v) uncertain, (vi) unknown phase, (vii) newly homozygous, or (viii) newly hemizygous. Definitions for newly homozygous, newly hemizygous, unknown phase, and uncertain inheritance types are available in our previous manuscript (Ruzzo et al. 2019). All VQSR failed and multi-allelic variants were excluded before reaching this classification step. Variants were also required to have a read depth of ≥10X, genotype quality of ≥25, and alternative allele reads/total reads ratio of ≥0.2.

#### Defining rare inherited and private variants

Rare inherited variants were defined as SNVs and indels with an allele frequency (AF) ≤0.1% among publicly available databases (1000g2015aug_all, ExACv3.0, cg46, gnomAD), internal controls, and iHART HNPs. These variants were also restricted to those not flagged as low-confidence in the Genome in a Bottle Consortium (GIAB; Zook et al., 2014) and missing in less than or equal to 25% of controls.

Private variants were defined as SNVs and indels observed in only one iHART family (AF∼0.05%) and missing in less than or equal to 25% of iHART HNPs. In addition, private variants were required to meet the following criteria: (i) not observed among control cohorts (AF=0), (ii) missing in less than or equal to 25% of PSP controls, (iii) not flagged as low-confidence in the GIAB Consortium. We only analyzed private variants among the 2,254 fully-phaseable children in our study.

#### Non-coding analyses

##### Definition of non-coding variants

In this manuscript, we defined non-coding SNVs and indels as variants that do not occur within a VEP annotated coding transcript. These include 17 of the 35 VEP consequences: “3_prime_UTR_variant”, “5_prime_UTR_variant”, “downstream_gene_variant”, “upstream_gene_variant”, “feature_elongation”, “feature_truncation”, “intergenic_variant”, “intron_variant”, “mature_miRNA_variant”, “non_coding_transcript_exon_variant”, “non_coding_transcript_variant”, “regulatory_region_ablation”, “regulatory_region_amplification”, “regulatory_region_variant”, “TF_binding_site_variant”, “TFBS_ablation”, “TFBS_amplification” (Ruzzo et al. 2019). To ensure strict filtering of non-coding variants, only the first most damaging consequence associated with a single variant were taken into account if there were multiple annotations available. We only considered non-coding variants that had not been flagged as low-confidence by the GIAB consortium.

##### Samples included for non-coding analyses

iHART non-coding rare and private inherited analyses included variants from 2,254 fully-phaseable children (N_aff_=1,836, N_unaff_=418). iHART non-coding RDNVs were restricted to 1,442 non-ARC outlier samples (N_aff_=1,164, N_unaff_=279) and variants identified as high confidence by ARC.

##### Description of non-coding variant sets tested

The non-coding rare inherited, private inherited, and rare *de novo* variant sets described above were further refined before quantitative burden testing, with the aim of increasing power to detect any association with ASD by narrowing variants to those more likely to be functional and/or contributing to the disorder according to previous studies. The first refined sets of non-coding variants used (Figure 4A-C) consist of those mapping to promoters, defined as both 2kb and 10kb upstream and 1kb downstream (3kb and 11kb promoters, respectively) from the transcription start site (TSS), by referencing the longest transcript for each gene (ties in transcript length were resolved by selecting the lower Ensembl Transcript ID). Rare inherited and private inherited variants mapping to 3kb promoters where further filtered (Figure S4D-E) by keeping those in promoters of genes belonging to ASD risk gene sets identified and defined in this study, the 74 TADA ASD risk genes (with FDR < 0.1 in TADA-mega analysis) and the 152 known ASD risk genes. We also specifically focused on variants mapping to transcription factor binding sites (TFBS) located within 11kb promoters (Figure S4F-H). As single global maps of genomic regions bound by transcription factors (TFs) to use for variant filtering we chose (1) cis-regulatory modules (CRMs) from the whole UniBind database (Gheorghe et al. 2019), and highly reliable (category A) (2) cistromes and (3) cismotifs, as defined in (Vorontsov et al. 2018). Finally, we took advantage of a new co-localization approach for interpreting the functionality of non-coding regions (Xu et al. 2020), which evaluates both their sequence constraint within the human lineage and their tissue- or cell type-specific regulatory role, by keeping variants in both 11kb promoters and across the whole genome that were mapping to the 10-tissue catalogue of brain-specific co-localized regions (http://www.funlda.com/colocalization/region) (Figure S4I-K).

##### Non-coding variant quantitative burden testing

Quantitative burden of non-coding variants in each refined set was compared between affected and unaffected children (Figure S4A-K) through both regression and other statistical tests (one-sample proportion test and Poisson rate ratio test) on total variant counts in the two groups. In addition, the relationship between phenotypic group (affected versus unaffected) and sex (female versus male) / sample source of DNA used for whole-genome sequencing (whole blood versus LCL) / carrying a non-coding rare *de novo* variant (carrier versus non-carrier) was explored by performing independence tests. All these statistical tests (p-values < 0.05 were considered significant) returned consistent and reproducible results. However, regression was the preferred approach for quantitative burden testing, since it gave us the advantage to correct for two important covariates, child sex and sample source of DNA used for whole-genome sequencing. Non-coding variants belonging to each refined set were counted for each child and used to perform regression, together with child phenotypic group, sex, and sample source of DNA. First, we modeled the response variable “variant count per child” by including the predictor variables “child phenotype”, “child sex”, and “sample source of DNA” through Poisson / quasi-Poisson / negative binomial regression models (R stats glm() function and MASS glm.nb() function were used). The negative binomial models were the best model to correct for over-dispersion of the count data. For rare *de novo* variants and both rare inherited and private inherited variants mapping to 3kb promoters of genes belonging to ASD risk gene sets (Figure S4D-E), we checked for potential zero-inflation of the count data and additionally built zero-inflated Poisson and zero-inflated negative binomial regression models (R pscl zeroinfl() function was used). Even for these variant sets, the negative binomial models were the best to predict zero counts. Alternatively, we modeled the response variable “child phenotype” by including the predictor variables “variant count per child”, “child sex”, and “sample source of DNA” through logistic regression. For all models built, we used (1) the residual deviance to perform a goodness of fit test for the overall model and (2) the difference between the residual deviance for the model with predictors and for the null model to test the significance of the overall model. These tests confirmed that the logistic regression models were the only models with a good fit and significance for the overall model. For this reason, although all approaches for quantitative burden testing described above returned consistent results, in this paper we chose to report those from logistic regression analyses (Table S2) for which we also computed power (Table S3). ASD odds ratio (OR) values were obtained by exponentiating the estimated regression coefficients.

##### Power analysis for quantitative burden testing through logistic regression

Power for quantitative burden testing through logistic regression was computed for each tested variant set by simulation. First, we used a posthoc approach, computing power for the observed ASD ORs and at the current sample sizes for all three predictor variables “variant count per child”, “child sex”, and “sample source of DNA”. We adopted three different simulation strategies to generate simulated data to use as new input to rerun logistic regression and estimate power, based on (1) the R simr powerSim() function (Green and MacLeod 2016) and on two custom functions named (2) “sample” and (3) “random”, respectively. The first two strategies are use bootstrapping, which resamples from the existing dataset, while the “random” strategy uses random variable generation and regression coefficients from the logistic model for the three predictor variables to compute the simulated response variable “child phenotype”. Quality of simulations was assessed by visualizing both simulated variables and resulting logistic regression coefficients. Power was estimated as the proportion of significant regression results (p < 0.05) over 1,000 simulations, together with 95% confidence intervals for this value. The three simulation strategies returned very consistent results (Table S3): in this paper we chose to report those obtained with the “sample” custom function strategy (Table S3 and Figures S4L-S4N). In addition, for each tested variant set we built power curves (through strategies 2 and 3, returning reproducible results) specific to the variant count per child predictor, for the observed ASD OR and at specific increasing sample sizes (private inherited: 2000-9000; rare *de novo*: 1,000-8,000). Again, power was estimated over 1,000 simulations. Curves were fitted by local regression for visualization.

The posthoc power analysis confirmed that the observed ASD ORs for the non-coding variant sets tested are too small to be detected at the current sample sizes, consistently with results from previous analysis of ASD simplex families (Werling et al. 2018) (Table S3). The power curves built for increasing sample sizes showed that a sample size of at least 8,000 fully-phaseable children would give us sufficient power to detect the observed ASD OR for private inherited variants mapping to 11kb promoter regions (at least 9,000 fully-phaseable children for the following private inherited sets: 3kb promoters, cistromes, cismotifs, and brain co-localized regions across the whole genome), whereas a sample size of at least 7,000 fully-phaseable children would allow us to detect the observed ASD OR for rare *de novo* variants mapping to brain co-localized regions across the whole genome (Figures S4M-N and Table S3 (as reference for ASD ORs and p values for each set)). Notably, the increasing sample sizes tested would not give us sufficient power to detect the observed ASD OR for any of the sets of rare inherited variants (Figure S4L and Table S3).

#### Artifact Removal by Classifier (ARC)

The Artifact Removal by Classifier (ARC) supervised model was designed to separate true rare *de novo* variants from LCL-specific genetic aberrations or other kinds of artifacts, such as sequencing and mapping errors (https://github.com/walllab/iHART-ARC). Details on the training, features, and performance evaluation of ARC are provided in our previous manuscript (Ruzzo et al. 2019).

##### ARC outlier samples

When applied to all 2,254 children (partially or fully-phaseable), we found a subset of outlier samples (fully-phaseable n=657) that had an ARC score less than 0.4 for >90% of their raw *de novo* variant calls. We excluded all ARC outlier samples from downstream analyses that involved *de novo* variants unless otherwise mentioned.

#### *De novo* variant rate vs. paternal age

We analyzed the correlation between paternal age and *de novo* variant rate in 1,157 fully-phaseable, affected iHART children (excluding MZ twins, ARC outliers, and 7 samples without paternal age information). Details on the linear model used in this analysis are available in our previous study (Ruzzo et al. 2019). We observed a robust signal after running ARC (*P*<2.2 x 10^-16^), but not prior to application of ARC (Figure S3D). According to our study, the RDNV rate increased by 1.02 per year of paternal age (95% CI = 1.02-1.03), consistent with previously published studies (Deciphering Developmental Disorders Study 2017; Francioli et al. 2015; Goldmann et al. 2016; Michaelson et al. 2012).

#### Rates for rare *de novo* variants

In our analysis, we only considered *de novo* variants in our 2,254 fully-phaseale children. Rare *de novo* variants were defined as: (i) absent in all controls, (ii) given an ARC score ≥0.4, (iii) missing in less than or equal to 25% of controls, (iv) no flagged as low-confidence by the GIAB consortium, (v) not present in an ARC outlier sample (n=657). Due to the use of LCL MZ twins (n=154) as the ARC training set, all MZ twin *de novo* variants were excluded from *de novo* rate calculations. Consequently, all *de novo* variant rate calculations were performed using 1,443 fully-phaseable non-MZ twins and non-ARC outliers (N_aff_=1,164; N_unaff_=279).

Our results suggest a mean genome-wide rate of 52.8 RDNVs per child (Figure S3B), which is consistent with previously reported rates (mean=64.4; range 54.8-81) (Besenbacher et al., 2016; Conrad et al., 2011; Kong et al., 2012; Michaelson et al., 2012; Turner et al., 2016; Yuen et al., 2017). We used the Wilcoxon rank sum test to determine if the number of *de novo* variants was significantly different in affected versus unaffected children.

#### Comparison of rates of variants in affected and unaffected children

For rare inherited, rare *de novo* and private inherited variants, we compared the rates (number of variants per child) for missense, synonymous and protein truncating variants in affected versus unaffected children. We used the logistic regression model:

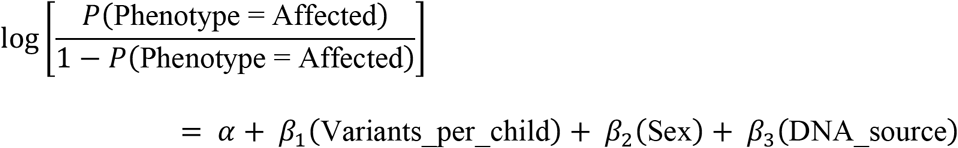

We performed this analysis with all genes and those with pLI>0.9 and 0.995

#### TADA mega-analysis

##### Samples and qualifying variants

In order to combine evidence found in ASD cases from rare *de novo* (DN) or transmitted (inherited) PTVs as well as *de novo* Mis3 variants predicted to damage the encoded protein, we used the Transmitted and *De novo* Association (TADA) test (He et al., 2013). Our TADA analysis was conducted on 1,660 genetically non-identical (one MZ twin retained) ASD cases from 864 iHART families with at least one ASD case and both biological parents sequenced. We treated these 1,660 affected children and their biological parents as independent trios in the TADA analysis. To enhance our ability to identify novel ASD-risk genes, we merged qualifying variants found in ASD cases from our current (iHART) cohort with a previous TADA mega-analysis (Sanders et al., 2015), which included both variants discovered from the Autism Sequencing Consortium (ASC) and Simons Simplex Collection (SSC), as well as small *de novo* CNV deletions (SmallDel) identified from SSC and Autism Genome Project (AGP) probands (Table S4).

Qualifying variants in our cohort included rare *de novo* and transmitted PTVs as well as *de novo* Mis3 variants present in the 1,660 affected samples, not flagged as low-confidence by the GIAB consortium (Zook et al., 2014). As in the previous TADA mega-analysis (Sanders et al., 2015), transmitted PTVs were required to have an AF≤0.1% among public databases (1000g, ESP6500, ExACv3.0, cg46, gnomAD), internal controls, and iHART HNPs. *De novo* variants were required to be absent (AF=0) across all public databases, internal controls, and iHART HNPs. ARC was run on all non-MZ twins to obtain high confidence *de novo* variants. *De novo* variants shared in MZ-twins were also considered qualifying variants and not filtered on their ARC score. For the TADA samples identified as ARC outliers (n=421), their *de novo* variants were excluded but their transmitted PTVs were retained as qualifying variants. We annotated the *de novo* Mis3 variants in the iHART cohort using the “probably damaging” PolyPhen-2 (Adzhubei et al., 2010) v2.2.2r395 HumDiv predictions from the Whole Human Exome Sequence Space (WHESS dataset). In cases where several qualifying variants in a gene were found in a sample, only the most damaging variant was retained.

For the TADA analysis, we then calculated the counts for all the qualifying variants from the different cohorts into a gene-by-variant-type matrix, which included variant counts for a total of 18,472 gencodeV19 genes with HGNC approved gene names. Specifically, DN PTV/Mis3 variants counts are identified in 5,275 ASD cases collected from iHART (this manuscript), SSC (Iossifov et al., 2014), and ASC (De Rubeis et al., 2014), while counts for the transmitted and non-transmitted PTVs are compiled from 4,563 ASD cases and 8,359 controls collected from the ASC (De Rubeis et al., 2014) and iHART (this manuscript) (Table S4). In addition, we calculated the counts of DN SmallDel in 4,687 ASD cases from the SSC (Sanders et al., 2015) and the AGP (Pinto et al., 2014) (Table S4).

We note that 119 of the 424 AGRE samples included as “cases” in the original TADA analysis (De Rubeis et al., 2014) with only transmitted PTVs accounted for, were able to have their DN PTV and Mis3 variants accounted for in iHART. This was due to iHART sequencing these samples (or their corresponding monozygotic twin) and their biological parents. The acquisition of these AGRE samples and the method used to count their DN PTV/Mis3 variants and transmitted PTVs are outlined in our previous manuscript (Ruzzo et al. 2019).

##### TADA parameters

The TADA parameters used in this study, match those used in our previous TADA mega-analysis (Ruzzo et al. 2019).

##### Novel gene discovery

For a gene to be considered novel, we applied strict parameters - it had to have an FDR <0.1 in our TADA mega-analysis and lack genome-wide statistical support across all previous studies (Sanders et al. 2015; Satterstrom et al. 2019; De Rubeis et al. 2014) with statistical rigor.

#### Genes with large inherited PTVs contribution

Given our signal for rare inherited variants, we sought to highlight genes for which a large contribution of the TADA ASD-risk association signal is derived from inherited PTVs. Conservatively, we considered only variants where the inheritance was known (*de novo* vs. inherited). Therefore, we adjusted the total number of TADA-qualifying variants to ignore PTVs from cases because some of the TADA-mega analysis qualifying variants originate from case-control studies (not iHART) where inheritance is unknown. We applied two methods to identify genes with a large contribution from inherited PTVs. The total number of qualifying variants (N) in each TADA gene was defined as N_DN.PTV_ + N_DN.SmallDel_ + N_DN.Mis3_ + N_Inherited.PTV_. We defined the percentage of inherited component of a gene as (N_Inherited.PTV_/N) x 100..

#### Fetal Single Cell Expression of iHART TADA analysis

Expression of iHART genes were examined in previously published single cell human developing neocortex, single cell fetal dataset (Polioudakis et al. 2019). The mean expression and percent expressed were calculated within each cell type and hierarchically clustered using base stats R function hclust() with “complete” method on the scaled values of the mean normalized expression within a cell type. To find cell type specific genes, the iHART genes were intersected with cell type enriched genes from (Polioudakis et al. 2019). Enriched genes from (Polioudakis et al. 2019) were found as follows: 1) wilcoxan rank sum test using FindAllMarkers from Seurat R package (Satija et al. 2015), 2) Bonferroni corrected p < 0.05, 3) greater than a 0.2 log fold change of expression in a given cluster compared to all other clusters and 4) detected in >10% of cells in a cell type cluster. Expression Weighted Cell Type Enrichment (EWCE) (Skene and Grant 2016) was performed to determine if TADA significant genes were enriched in cell types from (Polioudakis et al. 2019) using 10,000 bootstrap simulations and a background set of all coding genes measured in (Polioudakis et al. 2019).

#### Developmental Trajectory Analyses

Development trajectory RNAseq data was used from previous studies including the long term maturation human cortical spheroid (hCS) model (Gordon et al. 2021) and the BrainSpan dataset (Stein et al. 2014; Kang et al. 2011; M. Li et al. 2018). Briefly, hCS samples included six human induced pluripotent stem cell (hiPSC) lines derived from five different individuals that were cultured for up to 694 days *in vitro*. In total, 62 samples were collected for RNA sequencing (from four individuals, five hiPSC lines) at 13 timepoints. RNAseq libraries were prepared using Truseq stranded RNA RiboZero Gold (Illumina) and were sequenced using 100-bp paired end reads on an Illumina HiSeq 4000. Reads were then mapped to hg38 with Gencode v.25 annotations using STAR (v.2.5.2b) (Dobin et al. 2013). Gene expression levels were quantified using RSEM (v.1.3.0) (B. Li and Dewey 2011). Genes with low levels of expression (less than ten reads in more than 20% of the samples) were removed from the analysis. Outliers were then removed using standardized sample network connectivity (*Z* scores smaller than –3) (Oldham, Langfelder, and Horvath 2012). To quantify the technical variation in the RNA sequencing, we calculated the first five PCs of the Picard sequencing metrics (http://broadinstitute.github.io/picard/; v.2.5.0). These PCs, referred to as seqPC1–seqPC5, were then included in the linear model.

To help control for variability between the individuals racial background, we used the GATK (v.3.3) haplotype caller to call single nucleotide polymorphism (SNPs) from the aligned reads (McKenna et al. 2010). We filtered for sites with missing genotypes (>5%), rare minor allele frequency (<0.05) and out of Hardy–Weinberg equilibrium (<1×10^−6^) (Purcell et al. 2007). Genetic ancestry was inferred by running multidimensional scaling (MDS) on these high-quality SNPs together with HapMap3.3 (hg38). The first two MDS values, referred to as ancestryPC1/2, were then included in our linear model. For principal component analysis (PCA), as well as to visualize single gene trajectories, gene expression was normalized using CQN (without quantile normalization, sqn = FALSE) (v.1.28.0) and ancestryPC1-2 and SeqPC1-5 were regressed out before batch correction using Combat (Leek et al. 2012) from the sva package (v.3.30.0) in R. Single gene trajectories trends lines were fitted using the loess method (Cleveland and Devlin 1988) from the ggplot2 package (Wickham 2009, 2) in R.

In the BrainSpan RNA sequencing data (M. Li et al. 2018) to quantify gene expression at each developmental stage, the cortical samples were aligned to hg38 using Gencode v.25 annotations via STAR (Dobin et al. 2013). Gene expression was then quantified using the union exon model in featureCounts (Liao, Smyth, and Shi 2014). We removed low quality samples in which the RNA integrity number (RIN) was lower than 8, there were less than 25% coding bases or ribosomal bases made up more than 25% of total bases (as called by Picard tools). Genes with low levels of expression (less than ten mapped reads in more than 80% of the samples) in a given developmental stage were removed. We retained 196 samples from 24 individuals (9 female and 15 male).

We clustered the scaled normalized expression of hCS genes using hierarchical clustering on the Euclidean distance between genes. The most representative genes of a clustered were determined by each genes correlation with the cluster eigengene. Genes were clustered by their expression in the hCS using hierarchal clustering on the Euclidean distance between the genes. Cluster eigengenes were calculated using the module Eigengenes function from the WGCNA package (Langfelder and Horvath 2008). The gene in each cluster were correlated to the cluster module eigengene and the top five genes were annotated on the heatmap. The average expression per time point was calculated and trajectories trends lines were fitted using the loess method (Cleveland and Devlin 1988) from the ggplot2 package (Wickham 2009, 2). GO terms enrichment was performed using the enrichGO function from the clusterProfiler package (Yu et al. 2012) (v.3.12.0). Enrichment was performed on biological process and molecular function GO terms. All genes expressed in the hCS were used as background.

#### Known ASD Risk Genes (KARGs) Analysis

A comprehensive list of 152 known ASD-risk genes (KARG) was generated by merging unique genes, with HGNC approved gene names, identified at an FDR threshold of <0.1 in the iHART mega-analysis and previous TADA mega-analyses (Ruzzo et al., 2019; Sanders et al., 2015; Satterstrom et al., 2020). The same rare qualifying variants as TADA (DN/transmitted PTV and rare DN Mis3) were considered. Transmitted PTVs in ASD-risk genes were identified in the 2,254 fully-phaseable children (N_aff_=1,836, N_unaff_=418), while DN PTV or Mis3 qualifying variants in ASD-risk genes were restricted to those identified in the 1,443 non-ARC outlier samples (N_aff_=1,164, N_unaff_=279). We required transmitted PTVs to be in non-GIAB regions and have an AF≤0.1% in public databases (1000g, ESP6500, ExACv3.0, cg46, gnomAD), internal controls, and iHART HNP samples. DN variants identified in the iHART cohort were required to have an ARC score ≥0.4 and be in non-GIAB regions as well be absent in all public databases, internal controls, and HNP samples (AF = 0).

The list of risk genes for ASD and developmental disorders (DDD) was generated by merging unique genes, with HGNC approved gene names from known ASD risk genes and the Deciphering Development Disorders study (Deciphering Developmental Disorders Study 2017) with a FDR threshold of <0.1. This gene set totaled 208 genes (Table S5).

#### Polygenic Transmission Disequilibrium Test (pTDT) Analysis

##### Polygenic Risk Scoring

A polygenic risk score (PRS) was calculated to provide a quantitative estimate for an offspring’s genetic predisposition for ASD, SCZ, BD, and EA in iHART. To generate PRS, 3,255 samples of European ancestry, identified by multidimensional scaling (MDS)-derived hard race calls, were used (N_aff_=1,492, N_unaff_=1,763). For each disorder/trait tested, we obtained summary statistics from a GWAS with similar ancestry: for ASD we used the ASD iPSYCH GWAS (iHART overlaps with the PGC (Psychiatric Genomics Consortium) ASD GWAS; Naff=8,605, Nunaff=19,526) (Gandal et al. 2018); for SCZ we chose the SCZ CLOZUK+PGC meta-analysis GWAS (Naff= 40,675, Nunaff=64,643) (Pardiñas et al. 2018); for BD we used the BD PGC GWAS (Naff= 20,129, Nunaff=21,524) (Ruderfer et al. 2018); for EA (number of years of schooling completed) we chose the SSGAC (Social Science Genetic Association Consortium) EA meta-analysis GWAS (N= 293,723) (Okbay et al. 2016). Variants of interest were those in each GWAS with MAF≤0.05, mind≤0.1, geno=0, hwe≤1e-6. Candidate variant weights were then generated by running LDpred (Vilhjálmsson et al. 2015). PRS models corresponding to increasing fractions of contributing causal SNPs (0.001, 0.003, 0.01, 0.03, 0.1, 0.3, 1, Inf) were computed. Then, we evaluated the produced PRS models for each disorder/trait for their prediction of ASD affectation, using sex and the first two genotype principal components, PC1 and PC2, as model covariates. Sex is a confound of ASD affectation status. We removed (regressed out) PC1 and PC2 for the final evaluation of each PRS association with ASD affectation. In order to choose the best model for each disorder/trait, we examined both its Nagelkerke’s R2 and its regression coefficient for ASD affectation. Consistency and convergence for both measures across different p-value thresholds were observed at the p=1 one (assuming all SNPs are causal) that we chose uniformly for all phenotypes of interest. We observe a significant difference in ASD PRS between the tested iHART affected and unaffected subjects (p=0.01, beta=0.01, difference (vs. PRS null model) in Nagelkerke’s R2=0.003).

##### pTDT

We used the polygenic transmission disequilibrium test (pTDT) (Weiner et al. 2017) to determine whether an offspring’s genetic predisposition for ASD is consistent with what is expected based on parental transmission of polygenic risk for ASD. We first calculated the mid-parent polygenic risk score (PRS_MP_) for the 1,519 children of European ancestry, using the following equation:

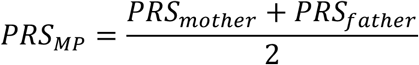

The PRS_MP_ was then used to compute the pTDT deviation for each child. The pTDT deviation is defined as follows:

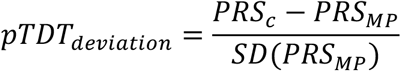

where PRS_C_ is the polygenic risk score for the affected or unaffected child, and SD(PRS_MP_) is the standard deviation of the sample-specific mid-parent PRS. In order to determine if the pTDT deviation was significantly different from zero, the two-sided, one-sample t-test was conducted using the pTDT test statistic (*t_pTDT_*):

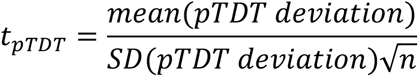

where *n* is the number of families included in the pTDT study. The same strategy was used for testing over-transmission of polygenic risk for SCZ, BD, and EA in iHART.

##### pTDT Variant Analysis

Carriers for variants in the 74 genes identified at an FDR threshold of <0.1 in the current iHART mega-analysis were considered in the pTDT analysis. The same rare qualifying variants as TADA (DN/transmitted PTV and rare DN Mis3) were considered. Transmitted PTVs in ASD-risk genes were identified in the 1,519 European children with both biological parents sequenced (fully-phaseable according to PRS calculation) who do not identify as a parent in the cohort as well (N_aff_=1,231, N_unaff_=288), while DN PTV or Mis3 qualifying variants in ASD-risk genes were restricted to those identified in the 1,066 non-ARC outlier samples (N_aff_=867, N_unaff_=199). We required transmitted PTVs to be in non-GIAB regions and have an AF≤0.1% in public databases (1000g, ESP6500, ExACv3.0, cg46, gnomAD), internal controls, and iHART HNP samples. DN variants identified in the iHART cohort were required to have an ARC score ≥0.4 and be in non-GIAB regions as well be absent in all public databases, internal controls, and HNP samples (AF = 0).

#### Phenotypic comparisons among children in iHART

We used phenotypic data available for iHART children to (1) investigate the phenotypic effect of rare inherited (RI) and rare *de novo* (RDN) variants in the 152 known ASD risk genes (KARG) and to (2) identify potential links between polygenic risk (PGR) over-transmission and phenotype severity through stratified pTDT based on children that do not carry any rare variant in the 74 TADA genes.

For the former analysis, we retrieved data available for the 2,254 fully-phaseable children whereas, for the latter one, we used those with phenotype available for the 1,519 fully-phaseable children of European ancestry.

We tested phenotypic measures descriptive of general intelligence, motor development, language development, and social behavior. The selected measures were verbal and non-verbal IQ scores, age of walking (AOW), ages of first word and phrase, and scores representing social behavior and social interaction skills. We used the standard scores from PPVT-3 (Peabody Picture Vocabulary Test, 3rd edition) (Dunn and Dunn 1997) and the Raven test (Raven and Raven 2003) as measures of verbal and non-verbal IQ, respectively. Age of walking and ages of first word and phrases in months were retrieved from ADI-R (Autism Diagnostic Interview-Revised, 2003 version with 93 items) (Rutter, Le Couteur, and Lord 2013). We made use of total raw scores and T-scores (with scoring by child’s sex) from SRS (Social Responsiveness Scale) (Constantino and Gruber 2005) as measures of deficiencies in social behavior.

For each measure, we used (1) the corresponding standard hypothesized value to which we compared the phenotypic distribution median value of each group of affected children (stratified by those carrying RI or RDN variants in KARG and those without such variants) and (2) cut-off value(s) for stratified pTDT analysis of affected TADA ASD risk gene non-carriers. IQ scores are normally distributed with a mean of 100 and a standard deviation of 15. We used the median value of 100 as the standard hypothesized value and a cut-off value of 70 (two standard deviations below the mean) to stratify children by those with (IQ < 70) or without (IQ >= 70) cognitive impairment.

To represent normal variation in age of walking, we used the window of achievement for the “walking alone” milestone observed in the normative cohort of the WHO Multicentre Growth Reference Study (MGRS) (Onis 2006). Specifically, the 50th percentile estimate of 12 months derived from this window was used as the standard hypothesized value, whereas the 97th percentile estimate of 16 months was used to divide children in those with atypical (AOW >= 16) or typical (AOW < 16) motor development.

Unaffected children from iHART were assessed for their language development through the AGRE (Lajonchere and AGRE Consortium 2010) custom seven-item language questionnaire. For both language measures, ages of first word and phrase, we used the available data to build “control non-carrier” distributions representing normal variation in milestone achievement among the unaffected siblings. We used the median values of 12 months and 18 months as reference hypothesized values for age of first word and age of first phrase, respectively. Both ADI-R items, age of first word and age of first phrase, are scored in section D of the diagnostic algorithm, with a score of 1 indicating that the age of milestone achievement suggests that developmental concerns are present at or before 36 months and a score of 0 denoting that the age of milestone achievement falls within the range of normal variation. We used this scoring system to stratify children by those with atypical (score = 1) or typical (score = 0) language development.

Total raw scores from SRS were available for both affected and unaffected children in iHART. We used the scores available for the unaffected non-carriers to build a reference distribution representing normal variation in social behavior and social interaction skills. We used the median score of 19 derived from this distribution as reference hypothesized value for testing. We computed corresponding T-scores using means, standard deviations, and sample sizes from the child’s sex-based school-age norm tables reported in the SRS manual (Constantino and Gruber 2012). We used the following SRS T-score cut-off ranges to divide the affected non-carriers into groups showing different degrees of social impairment: T-scores equal to 59 or less were considered as normative, those among 60 and 75 as representative of mild ASD, and those equal to 76 or more as characteristic of severe ASD (Lyall et al. 2014).

When available, we compared redundant measures for the same phenotype derived from different assessment tools to check general quality and concordance of the phenotypic data (not shown).

When data were collected using different version forms for the same assessment tool, we only used those coming from the most frequently used version form to ensure that form version did not drive potential phenotypic differences. An exception was made for SRS, for which we combined the compatible preschool and school-age forms.

When data for the same child were collected more than once, in more than one visit, we kept only those from the most recent visit for further analysis. Again, a different criterium was used for SRS. We removed data from assessments flagged as invalid. When valid SRS data for the same child were collected in more than one visit, (1) if both a parent and a teacher were available as corresponding respondents, the teacher-based assessment was preferred as more reliable, whereas (2) if the same respondent contributed to longitudinal assessments, the earliest visit data was used for downstream analysis (Lowe et al. 2015). In addition, since SRS has not been validated in non-verbal children, we used the ADI-R item age of first word to identify children that had not reached or regained this milestone and we removed their entries from the SRS dataset (Lowe et al. 2015). Finally, we computed sex-based T-scores only for affected non-carriers with ages 4 to 18 years (school-age sample).

Many children in the iHART cohort were still not able to walk or were non-verbal at the time of testing. We wanted to still keep these informative observations for the analysis despite not having a numeric value for those cases. Therefore, we converted the codes for “milestone not reached” and “milestone lost and not regained yet” into the age of those children at the time of testing plus one month, optimistically assuming for them a potential milestone achievement soon after the latest assessment. We verified that this approach did not remarkably alter the distributions observed for the three items through comparison with those built from the smaller dataset in which the code cases had been simply removed (not shown). Similarly, with the aim of not losing relevant information coming from our Raven data, we converted the codes for “below age-specific test norms” and “above age-specific test norms” non-verbal IQ scores into the cohort lowest minus one point and highest plus one point scores, respectively. There were two cases of affected “double carrier” of both a RDN and a RI variant in KARG and a case of affected “double carrier” of variants in the TADA ASD risk genes. We decided to group these children as RDN carriers, assuming a larger effect size for this rare genetic variation type.

We excluded some specific groups of children from these phenotypic comparison analyses: affected children with confirmed motor development impairment (cerebral palsy diagnosis), affected children with seizures, unaffected children with learning disabilities, unaffected children with language disabilities, unaffected children with previous and no longer confirmed ASD diagnosis, and unaffected children carrying qualifying rare variants in KARG and in the TADA ASD risk genes. These children were removed with the idea of minimizing the potential confounding effect of comorbid phenotypes and ASD-associated genetic variation on the measures tested. Even though we had IQ scores available for very few unaffected children we decided to remove them from the analyses, keeping them only on affected children. We only used data from unaffected children to build the ADI-R item and SRS total raw score “control non-carrier” distributions mentioned above.

Since the phenotypic measure distributions were not normal and asymmetric, we tested potential differences between their median values and standard hypothesized values through two-sided one-sample sign tests. The three tests performed for each phenotypic measure were considered as simultaneous multiple tests and we used the Benjamini and Hochberg (BH) procedure as FDR (false discovery rate)-controlling method. An adjusted p-value < 0.05 was considered significant. For the phenotypic measure-stratified pTDT analysis, we compared the pTDT deviation distribution mean values against the expected value of 0 through two-sided one-sample t-tests. We defined a nominal p-value < 0.05 as significant.

### QUANTIFICATION AND STATISTICAL ANALYSIS

Statistics were calculated using R (3.5.1) unless otherwise notes. The significance of PPI networks were evaluated using DAPPLE metrics based on 1,000 permutations (within the DAPPLE parameter); P values > 0.05 were considered significant.

#### TADA

The sample sizes for the TADA-mega analysis are provided in Table S4. After performing the Benjamini-Hochberg correction on the TADA results, q-values (False Discovery Rate (FDR)) <0.1 were considered significantly associated with ASD. We find 74 genome-wide significant genes when we apply the field standard FDR<0.1.

### DATA AND CODE AVAILABILITY

Following request and approval of the data use agreement found at http://www.ihart.org, the whole-genome sequencing data generated during this study can be downloaded from the Autism Research and Technology Initiative (iHART) of the Hartwell Foundation. Autism Speaks and AGRE must approve the release of whole-genome sequencing data generated during this study.

We provide the source code and a full tutorial for ARC (Artifact Removal by Classifier), a random forest supervised model developed to distinguish rare, *de novo* variants from LCL-specific genetic aberrations or other types of artifacts such as sequencing and mapping errors, at https://github.com/walllab/iHART-ARC.

## Notes

### Competing Interest Statement

The authors have declared no competing interest.

### Funding Statement

This study was funded by grants from The Hartwell Foundation and the NIH (R01MH100027, R01MH064547, S10OD011939, P50HD055784, UM1HG008901, P50DC018006, K08AG065519 R01MH064547, S10OD011939) and from the Stanford Precision Health and Integrated Diagnostics Center and from the Stanford Bio-X Center.

### Author Declarations

Ethics committee/IRB of University of California, Los Angeles waived ethical approval for this work Ethics committee/IRB of Stanford Univerity waived ethical approval for this work

## References

1000 Genomes Project Consortium, Adam Auton, Lisa D. Brooks, Richard M. Durbin, Erik P. Garrison, Hyun Min Kang, Jan O. Korbel, et al. 2015. “A Global Reference for Human Genetic Variation.” Nature 526 (7571): 68–74. https://doi.org/10.1038/nature15393.

Abbott, Angela E, Annika C Linke, Aarti Nair, Afrooz Jahedi, Laura A Alba, Christopher L Keown, Inna Fishman, and Ralph-Axel Müller. 2018. “Repetitive Behaviors in Autism Are Linked to Imbalance of Corticostriatal Connectivity: A Functional Connectivity MRI Study.” Social Cognitive and Affective Neuroscience 13 (1): 32–42. https://doi.org/10.1093/scan/nsx129.

Abrahams, Brett S., and Daniel H. Geschwind. 2008. “Advances in Autism Genetics: On the Threshold of a New Neurobiology.” Nature Reviews. Genetics 9 (5): 341–55. https://doi.org/10.1038/nrg2346.

Adzhubei, Ivan A., Steffen Schmidt, Leonid Peshkin, Vasily E. Ramensky, Anna Gerasimova, Peer Bork, Alexey S. Kondrashov, and Shamil R. Sunyaev. 2010. “A Method and Server for Predicting Damaging Missense Mutations.” Nature Methods 7 (4): 248–49. https://doi.org/10.1038/nmeth0410-248.

An, Joon-Yong, Kevin Lin, Lingxue Zhu, Donna M. Werling, Shan Dong, Harrison Brand, Harold Z. Wang, et al. 2018. “Genome-Wide de Novo Risk Score Implicates Promoter Variation in Autism Spectrum Disorder.” Science (New York, N.Y.) 362 (6420). https://doi.org/10.1126/science.aat6576.

Antaki, D., A. Maihofer, M. Klein, J. Guevara, J. Grove, Caitlin Carey, O. Hong, et al. 2021. “A Phenotypic Spectrum of Autism Is Attributable to the Combined Effects of Rare Variants, Polygenic Risk and Sex.” https://doi.org/10.1101/2021.03.30.21254657.

Bai, Dan, Benjamin Hon Kei Yip, Gayle C. Windham, Andre Sourander, Richard Francis, Rinat Yoffe, Emma Glasson, et al. 2019. “Association of Genetic and Environmental Factors With Autism in a 5-Country Cohort.” JAMA Psychiatry 76 (10): 1035–43. https://doi.org/10.1001/jamapsychiatry.2019.1411.

Barnett, Derek W., Erik K. Garrison, Aaron R. Quinlan, Michael P. Strömberg, and Gabor T. Marth. 2011. “BamTools: A C++ API and Toolkit for Analyzing and Managing BAM Files.” Bioinformatics (Oxford, England) 27 (12): 1691–92. https://doi.org/10.1093/bioinformatics/btr174.

Bell, L., A. Wittkowski, and D. J. Hare. 2019. “Movement Disorders and Syndromic Autism: A Systematic Review.” Journal of Autism and Developmental Disorders 49 (1): 54–67. https://doi.org/10.1007/s10803-018-3658-y.

Besenbacher, Søren, Patrick Sulem, Agnar Helgason, Hannes Helgason, Helgi Kristjansson, Aslaug Jonasdottir, Adalbjorg Jonasdottir, et al. 2016. “Multi-Nucleotide de Novo Mutations in Humans.” PLoS Genetics 12 (11): e1006315. https://doi.org/10.1371/journal.pgen.1006315.

Brandler, William M., Danny Antaki, Madhusudan Gujral, Morgan L. Kleiber, Joe Whitney, Michelle S. Maile, Oanh Hong, et al. 2018. “Paternally Inherited Cis-Regulatory Structural Variants Are Associated with Autism.” Science (New York, N.Y.) 360 (6386): 327–31. https://doi.org/10.1126/science.aan2261.

Bucan, Maja, Brett S. Abrahams, Kai Wang, Joseph T. Glessner, Edward I. Herman, Lisa I. Sonnenblick, Ana I. Alvarez Retuerto, et al. 2009. “Genome-Wide Analyses of Exonic Copy Number Variants in a Family-Based Study Point to Novel Autism Susceptibility Genes.” PLoS Genetics 5 (6): e1000536. https://doi.org/10.1371/journal.pgen.1000536.

Chung, K. K., Y. Zhang, K. L. Lim, Y. Tanaka, H. Huang, J. Gao, C. A. Ross, V. L. Dawson, and T. M. Dawson. 2001. “Parkin Ubiquitinates the Alpha-Synuclein-Interacting Protein, Synphilin-1: Implications for Lewy-Body Formation in Parkinson Disease.” Nature Medicine 7 (10): 1144–50. https://doi.org/10.1038/nm1001-1144.

Cleveland, William S., and Susan J. Devlin. 1988. “Locally Weighted Regression: An Approach to Regression Analysis by Local Fitting.” Journal of the American Statistical Association 83 (403): 596–610. https://doi.org/10.1080/01621459.1988.10478639.

Coe, Bradley P., Holly A.F. Stessman, Arvis Sulovari, Madeleine R. Geisheker, Trygve E. Bakken, Allison M. Lake, Joseph D. Dougherty, et al. 2019. “Neurodevelopmental Disease Genes Implicated by de Novo Mutation and Copy Number Variation Morbidity.” Nature Genetics 51 (1): 106–16. https://doi.org/10.1038/s41588-018-0288-4.

Colvert, Emma, Beata Tick, Fiona McEwen, Catherine Stewart, Sarah R. Curran, Emma Woodhouse, Nicola Gillan, et al. 2015. “Heritability of Autism Spectrum Disorder in a UK Population-Based Twin Sample.” JAMA Psychiatry 72 (5): 415–23. https://doi.org/10.1001/jamapsychiatry.2014.3028.

Conrad, Donald F., Jonathan E. M. Keebler, Mark A. DePristo, Sarah J. Lindsay, Yujun Zhang, Ferran Casals, Youssef Idaghdour, et al. 2011. “Variation in Genome-Wide Mutation Rates within and between Human Families.” Nature Genetics 43 (7): 712–14. https://doi.org/10.1038/ng.862.

Constantino, John N., and C. Gruber. 2005. The Social Responsiveness Scale (SRS). Los Angeles, CA: Western Psychological Services.

Constantino, John N., and C. Gruber. 2012. Social Responsiveness Scale, Second Edition (SRS-2). Torrance, CA: Western Psychological Services.

Cross-Disorder Group of the Psychiatric Genomics Consortium. 2019. “Genomic Relationships, Novel Loci, and Pleiotropic Mechanisms across Eight Psychiatric Disorders.” Cell 179 (7): 1469–1482.e11. https://doi.org/10.1016/j.cell.2019.11.020.

Dashtipour, Khashayar, Ali Tafreshi, Charles Adler, Thomas Beach, Xin Chen, Geidy Serrano, Stephanie Tashiro, and Charles Wang. 2017. “Hypermethylation of Synphilin-1, Alpha-Synuclein-Interacting Protein (SNCAIP) Gene in the Cerebral Cortex of Patients with Sporadic Parkinson’s Disease.” Brain Sciences 7 (7). https://doi.org/10.3390/brainsci7070074.

De Rubeis, Silvia, Xin He, Arthur P. Goldberg, Christopher S. Poultney, Kaitlin Samocha, A. Erucment Cicek, Yan Kou, et al. 2014. “Synaptic, Transcriptional and Chromatin Genes Disrupted in Autism.” Nature 515 (7526): 209–15. https://doi.org/10.1038/nature13772.

Deciphering Developmental Disorders Study. 2017. “Prevalence and Architecture of de Novo Mutations in Developmental Disorders.” Nature 542 (7642): 433–38. https://doi.org/10.1038/nature21062.

DePristo, Mark A., Eric Banks, Ryan Poplin, Kiran V. Garimella, Jared R. Maguire, Christopher Hartl, Anthony A. Philippakis, et al. 2011. “A Framework for Variation Discovery and Genotyping Using Next-Generation DNA Sequencing Data.” Nature Genetics 43 (5): 491–98. https://doi.org/10.1038/ng.806.

Dobin, Alexander, Carrie A. Davis, Felix Schlesinger, Jorg Drenkow, Chris Zaleski, Sonali Jha, Philippe Batut, Mark Chaisson, and Thomas R. Gingeras. 2013. “STAR: Ultrafast Universal RNA-Seq Aligner.” Bioinformatics (Oxford, England) 29 (1): 15–21. https://doi.org/10.1093/bioinformatics/bts635.

Dong, Shan, Michael F. Walker, Nicholas J. Carriero, Michael DiCola, A. Jeremy Willsey, Adam Y. Ye, Zainulabedin Waqar, et al. 2014. “De Novo Insertions and Deletions of Predominantly Paternal Origin Are Associated with Autism Spectrum Disorder.” Cell Reports 9 (1): 16–23. https://doi.org/10.1016/j.celrep.2014.08.068.

Drmanac, Radoje, Andrew B. Sparks, Matthew J. Callow, Aaron L. Halpern, Norman L. Burns, Bahram G. Kermani, Paolo Carnevali, et al. 2010. “Human Genome Sequencing Using Unchained Base Reads on Self-Assembling DNA Nanoarrays.” Science (New York, N.Y.) 327 (5961): 78–81. https://doi.org/10.1126/science.1181498.

Dunn, L.M., and L.M. Dunn. 1997. Peabody Picture Vocabulary Test-III. Circle Pines, MN: American Guidance Service (AGS).

Engelender, S., Z. Kaminsky, X. Guo, A. H. Sharp, R. K. Amaravi, J. J. Kleiderlein, R. L. Margolis, et al. 1999. “Synphilin-1 Associates with Alpha-Synuclein and Promotes the Formation of Cytosolic Inclusions.” Nature Genetics 22 (1): 110–14. https://doi.org/10.1038/8820.

Epi4K Consortium, Epilepsy Phenome/Genome Project, Andrew S. Allen, Samuel F. Berkovic, Patrick Cossette, Norman Delanty, Dennis Dlugos, et al. 2013. “De Novo Mutations in Epileptic Encephalopathies.” Nature 501 (7466): 217–21. https://doi.org/10.1038/nature12439.

Estes, Annette, Dennis W. W. Shaw, Bobbi F. Sparks, Seth Friedman, Jay N. Giedd, Geraldine Dawson, Matthew Bryan, and Stephen R. Dager. 2011. “Basal Ganglia Morphometry and Repetitive Behavior in Young Children with Autism Spectrum Disorder.” Autism Research: Official Journal of the International Society for Autism Research 4 (3): 212– 20. https://doi.org/10.1002/aur.193.

Feliciano, Pamela, Xueya Zhou, Irina Astrovskaya, Tychele N. Turner, Tianyun Wang, Leo Brueggeman, Rebecca Barnard, et al. 2019. “Exome Sequencing of 457 Autism Families Recruited Online Provides Evidence for Autism Risk Genes.” NPJ Genomic Medicine 4: 19. https://doi.org/10.1038/s41525-019-0093-8.

Fombonne, Eric. 2009. “Epidemiology of Pervasive Developmental Disorders.” Pediatric Research 65 (6): 591–98. https://doi.org/10.1203/PDR.0b013e31819e7203.

Francioli, Laurent C., Paz P. Polak, Amnon Koren, Androniki Menelaou, Sung Chun, Ivo Renkens, Cornelia M. van Duijn, et al. 2015. “Genome-Wide Patterns and Properties of de Novo Mutations in Humans.” Nature Genetics 47 (7): 822–26. https://doi.org/10.1038/ng.3292.

Gandal, Michael J., Jillian R. Haney, Neelroop N. Parikshak, Virpi Leppa, Gokul Ramaswami, Chris Hartl, Andrew J. Schork, et al. 2018. “Shared Molecular Neuropathology across Major Psychiatric Disorders Parallels Polygenic Overlap.” Science (New York, N.Y.) 359 (6376): 693–97. https://doi.org/10.1126/science.aad6469.

Gaugler, Trent, Lambertus Klei, Stephan J. Sanders, Corneliu A. Bodea, Arthur P. Goldberg, Ann B. Lee, Milind Mahajan, et al. 2014. “Most Genetic Risk for Autism Resides with Common Variation.” Nature Genetics 46 (8): 881–85. https://doi.org/10.1038/ng.3039.

Geschwind, Daniel H., J. Sowinski, C. Lord, P. Iversen, J. Shestack, P. Jones, L. Ducat, S. J. Spence, and AGRE Steering Committee. 2001. “The Autism Genetic Resource Exchange: A Resource for the Study of Autism and Related Neuropsychiatric Conditions.” American Journal of Human Genetics 69 (2): 463–66. https://doi.org/10.1086/321292.

Geschwind, Daniel H., and Matthew W. State. 2015. “Gene Hunting in Autism Spectrum Disorder: On the Path to Precision Medicine.” The Lancet. Neurology 14 (11): 1109–20. https://doi.org/10.1016/S1474-4422(15)00044-7.

Gheorghe, Marius, Geir Kjetil Sandve, Aziz Khan, Jeanne Chèneby, Benoit Ballester, and Anthony Mathelier. 2019. “A Map of Direct TF-DNA Interactions in the Human Genome.” Nucleic Acids Research 47 (4): e21. https://doi.org/10.1093/nar/gky1210.

Goldmann, Jakob M., Wendy S. W. Wong, Michele Pinelli, Terry Farrah, Dale Bodian, Anna B. Stittrich, Gustavo Glusman, et al. 2016. “Parent-of-Origin-Specific Signatures of de Novo Mutations.” Nature Genetics 48 (8): 935–39. https://doi.org/10.1038/ng.3597.

Gordon, Aaron, Se-Jin Yoon, Stephen S. Tran, Christopher D. Makinson, Jin Young Park, Jimena Andersen, Alfredo M. Valencia, et al. 2021. “Long-Term Maturation of Human Cortical Organoids Matches Key Early Postnatal Transitions.” Nature Neuroscience 24 (3): 331–42. https://doi.org/10.1038/s41593-021-00802-y.

Green, Peter, and Catriona J. MacLeod. 2016. “SIMR: An R Package for Power Analysis of Generalized Linear Mixed Models by Simulation.” Methods in Ecology and Evolution 7 (4): 493–98. https://doi.org/10.1111/2041-210X.12504.

Grove, Jakob, Stephan Ripke, Thomas D. Als, Manuel Mattheisen, Raymond K. Walters, Hyejung Won, Jonatan Pallesen, et al. 2019. “Identification of Common Genetic Risk Variants for Autism Spectrum Disorder.” Nature Genetics 51 (3): 431–44. https://doi.org/10.1038/s41588-019-0344-8.

He, Xin, Stephan J. Sanders, Li Liu, Silvia De Rubeis, Elaine T. Lim, James S. Sutcliffe, Gerard D. Schellenberg, et al. 2013. “Integrated Model of de Novo and Inherited Genetic Variants Yields Greater Power to Identify Risk Genes.” PLoS Genetics 9 (8): e1003671. https://doi.org/10.1371/journal.pgen.1003671.

Hu, Youna, Alena Shmygelska, David Tran, Nicholas Eriksson, Joyce Y. Tung, and David A. Hinds. 2016. “GWAS of 89,283 Individuals Identifies Genetic Variants Associated with Self-Reporting of Being a Morning Person.” Nature Communications 7 (February): 10448. https://doi.org/10.1038/ncomms10448.

Iossifov, Ivan, Brian J. O’Roak, Stephan J. Sanders, Michael Ronemus, Niklas Krumm, Dan Levy, Holly A. Stessman, et al. 2014. “The Contribution of de Novo Coding Mutations to Autism Spectrum Disorder.” Nature 515 (7526): 216–21. https://doi.org/10.1038/nature13908.

Jacquemont, Sébastien, Bradley P. Coe, Micha Hersch, Michael H. Duyzend, Niklas Krumm, Sven Bergmann, Jacques S. Beckmann, Jill A. Rosenfeld, and Evan E. Eichler. 2014. “A Higher Mutational Burden in Females Supports a ‘Female Protective Model’ in Neurodevelopmental Disorders.” American Journal of Human Genetics 94 (3): 415–25. https://doi.org/10.1016/j.ajhg.2014.02.001.

Jun, Goo, Matthew Flickinger, Kurt N. Hetrick, Jane M. Romm, Kimberly F. Doheny, Gonçalo R. Abecasis, Michael Boehnke, and Hyun Min Kang. 2012. “Detecting and Estimating Contamination of Human DNA Samples in Sequencing and Array-Based Genotype Data.” American Journal of Human Genetics 91 (5): 839–48. https://doi.org/10.1016/j.ajhg.2012.09.004.

Kang, Hyo Jung, Yuka Imamura Kawasawa, Feng Cheng, Ying Zhu, Xuming Xu, Mingfeng Li, André M. M. Sousa, et al. 2011. “Spatio-Temporal Transcriptome of the Human Brain.” Nature 478 (7370): 483–89. https://doi.org/10.1038/nature10523.

Karczewski, Konrad J., Laurent C. Francioli, Grace Tiao, Beryl B. Cummings, Jessica Alföldi, Qingbo Wang, Ryan L. Collins, et al. 2020. “The Mutational Constraint Spectrum Quantified from Variation in 141,456 Humans.” BioRxiv, April, 531210. https://doi.org/10.1101/531210.

Kong, Augustine, Michael L. Frigge, Gisli Masson, Soren Besenbacher, Patrick Sulem, Gisli Magnusson, Sigurjon A. Gudjonsson, et al. 2012. “Rate of de Novo Mutations and the Importance of Father’s Age to Disease Risk.” Nature 488 (7412): 471–75. https://doi.org/10.1038/nature11396.

Krüger, Rejko. 2004. “The Role of Synphilin-1 in Synaptic Function and Protein Degradation.” Cell and Tissue Research 318 (1): 195–99. https://doi.org/10.1007/s00441-004-0953-z.

Krumm, Niklas, Tychele N. Turner, Carl Baker, Laura Vives, Kiana Mohajeri, Kali Witherspoon, Archana Raja, et al. 2015. “Excess of Rare, Inherited Truncating Mutations in Autism.” Nature Genetics 47 (6): 582–88. https://doi.org/10.1038/ng.3303.

Lajonchere, Clara M. and AGRE Consortium. 2010. “Changing the Landscape of Autism Research: The Autism Genetic Resource Exchange.” Neuron 68 (2): 187–91. https://doi.org/10.1016/j.neuron.2010.10.009.

Lane, Jacqueline M., Irma Vlasac, Simon G. Anderson, Simon D. Kyle, William G. Dixon, David A. Bechtold, Shubhroz Gill, et al. 2016. “Genome-Wide Association Analysis Identifies Novel Loci for Chronotype in 100,420 Individuals from the UK Biobank.” Nature Communications 7 (March): 10889. https://doi.org/10.1038/ncomms10889.

Langfelder, Peter, and Steve Horvath. 2008. “WGCNA: An R Package for Weighted Correlation Network Analysis.” BMC Bioinformatics 9 (December): 559. https://doi.org/10.1186/1471-2105-9-559.

Leek, Jeffrey T., W. Evan Johnson, Hilary S. Parker, Andrew E. Jaffe, and John D. Storey. 2012. “The Sva Package for Removing Batch Effects and Other Unwanted Variation in High-Throughput Experiments.” Bioinformatics (Oxford, England) 28 (6): 882–83. https://doi.org/10.1093/bioinformatics/bts034.

Lek, Monkol, Konrad J. Karczewski, Eric V. Minikel, Kaitlin E. Samocha, Eric Banks, Timothy Fennell, Anne H. O’Donnell-Luria, et al. 2016. “Analysis of Protein-Coding Genetic Variation in 60,706 Humans.” Nature 536 (7616): 285–91. https://doi.org/10.1038/nature19057.

Leppa, Virpi M., Stephanie N. Kravitz, Christa Lese Martin, Joris Andrieux, Cedric Le Caignec, Dominique Martin-Coignard, Christina DyBuncio, et al. 2016. “Rare Inherited and De Novo CNVs Reveal Complex Contributions to ASD Risk in Multiplex Families.” American Journal of Human Genetics 99 (3): 540–54. https://doi.org/10.1016/j.ajhg.2016.06.036.

Li, Bo, and Colin N. Dewey. 2011. “RSEM: Accurate Transcript Quantification from RNA-Seq Data with or without a Reference Genome.” BMC Bioinformatics 12 (August): 323. https://doi.org/10.1186/1471-2105-12-323.

Li, Heng, Bob Handsaker, Alec Wysoker, Tim Fennell, Jue Ruan, Nils Homer, Gabor Marth, Goncalo Abecasis, Richard Durbin, and 1000 Genome Project Data Processing Subgroup. 2009. “The Sequence Alignment/Map Format and SAMtools.” Bioinformatics (Oxford, England) 25 (16): 2078–79. https://doi.org/10.1093/bioinformatics/btp352.

Li, Mingfeng, Gabriel Santpere, Yuka Imamura Kawasawa, Oleg V. Evgrafov, Forrest O. Gulden, Sirisha Pochareddy, Susan M. Sunkin, et al. 2018. “Integrative Functional Genomic Analysis of Human Brain Development and Neuropsychiatric Risks.” Science (New York, N.Y.) 362 (6420). https://doi.org/10.1126/science.aat7615.

Liao, Yang, Gordon K. Smyth, and Wei Shi. 2014. “FeatureCounts: An Efficient General Purpose Program for Assigning Sequence Reads to Genomic Features.” Bioinformatics (Oxford, England) 30 (7): 923–30. https://doi.org/10.1093/bioinformatics/btt656.

Lord, C., S. Risi, L. Lambrecht, E. H. Cook, B. L. Leventhal, P. C. DiLavore, A. Pickles, and M. Rutter. 2000. “The Autism Diagnostic Observation Schedule-Generic: A Standard Measure of Social and Communication Deficits Associated with the Spectrum of Autism.” Journal of Autism and Developmental Disorders 30 (3): 205–23.

Lord, C., M. Rutter, and A. Le Couteur. 1994. “Autism Diagnostic Interview-Revised: A Revised Version of a Diagnostic Interview for Caregivers of Individuals with Possible Pervasive Developmental Disorders.” Journal of Autism and Developmental Disorders 24 (5): 659–85. https://doi.org/10.1007/BF02172145.

Lowe, Jennifer K., Donna M. Werling, John N. Constantino, Rita M. Cantor, and Daniel H. Geschwind. 2015. “Social Responsiveness, an Autism Endophenotype: Genomewide Significant Linkage to Two Regions on Chromosome 8.” The American Journal of Psychiatry 172 (3): 266–75. https://doi.org/10.1176/appi.ajp.2014.14050576.

Lyall, Kristen, John N. Constantino, Marc G. Weisskopf, Andrea L. Roberts, Alberto Ascherio, and Susan L. Santangelo. 2014. “Parental Social Responsiveness and Risk of Autism Spectrum Disorder in Offspring.” JAMA Psychiatry 71 (8): 936–42. https://doi.org/10.1001/jamapsychiatry.2014.476.

Maenner, Matthew J., Kelly A. Shaw, Jon Baio, EdS1, Anita Washington, Mary Patrick, Monica DiRienzo, et al. 2020. “Prevalence of Autism Spectrum Disorder Among Children Aged 8 Years - Autism and Developmental Disabilities Monitoring Network, 11 Sites, United States, 2016.” Morbidity and Mortality Weekly Report. Surveillance Summaries (Washington, D.C.: 2002) 69 (4): 1–12. https://doi.org/10.15585/mmwr.ss6904a1.

Marshall, Christian R., Abdul Noor, John B. Vincent, Anath C. Lionel, Lars Feuk, Jennifer Skaug, Mary Shago, et al. 2008. “Structural Variation of Chromosomes in Autism Spectrum Disorder.” American Journal of Human Genetics 82 (2): 477–88. https://doi.org/10.1016/j.ajhg.2007.12.009.

McKenna, Aaron, Matthew Hanna, Eric Banks, Andrey Sivachenko, Kristian Cibulskis, Andrew Kernytsky, Kiran Garimella, et al. 2010. “The Genome Analysis Toolkit: A MapReduce Framework for Analyzing next-Generation DNA Sequencing Data.” Genome Research 20 (9): 1297–1303. https://doi.org/10.1101/gr.107524.110.

McLaren, William, Laurent Gil, Sarah E. Hunt, Harpreet Singh Riat, Graham R. S. Ritchie, Anja Thormann, Paul Flicek, and Fiona Cunningham. 2016. “The Ensembl Variant Effect Predictor.” Genome Biology 17 (1): 122. https://doi.org/10.1186/s13059-016-0974-4.

Michaelson, Jacob J., Yujian Shi, Madhusudan Gujral, Hancheng Zheng, Dheeraj Malhotra, Xin Jin, Minghan Jian, et al. 2012. “Whole-Genome Sequencing in Autism Identifies Hot Spots for de Novo Germline Mutation.” Cell 151 (7): 1431–42. https://doi.org/10.1016/j.cell.2012.11.019.

Neale, Benjamin M., Yan Kou, Li Liu, Avi Ma’ayan, Kaitlin E. Samocha, Aniko Sabo, Chiao-Feng Lin, et al. 2012. “Patterns and Rates of Exonic de Novo Mutations in Autism Spectrum Disorders.” Nature 485 (7397): 242–45. https://doi.org/10.1038/nature11011.

O’Brien, Heath E., Eilis Hannon, Aaron R. Jeffries, William Davies, Matthew J. Hill, Richard J. Anney, Michael C. O’Donovan, Jonathan Mill, and Nicholas J. Bray. 2019. “Sex Differences in Gene Expression in the Human Fetal Brain.” *BioRxiv*, March, 483636. https://doi.org/10.1101/483636.

Okbay, Aysu, Jonathan P. Beauchamp, Mark Alan Fontana, James J. Lee, Tune H. Pers, Cornelius A. Rietveld, Patrick Turley, et al. 2016. “Genome-Wide Association Study Identifies 74 Loci Associated with Educational Attainment.” Nature 533 (7604): 539–42. https://doi.org/10.1038/nature17671.

Oldham, Michael C., Peter Langfelder, and Steve Horvath. 2012. “Network Methods for Describing Sample Relationships in Genomic Datasets: Application to Huntington’s Disease.” BMC Systems Biology 6 (June): 63. https://doi.org/10.1186/1752-0509-6-63.

Oliva, Meritxell, Manuel Muñoz-Aguirre, Sarah Kim-Hellmuth, Valentin Wucher, Ariel D. H. Gewirtz, Daniel J. Cotter, Princy Parsana, et al. 2020. “The Impact of Sex on Gene Expression across Human Tissues.” Science (New York, N.Y.) 369 (6509). https://doi.org/10.1126/science.aba3066.

Onis, Mercedes de. 2006. “WHO Motor Development Study: Windows of Achievement for Six Gross Motor Development Milestones.” Acta Paediatrica 95 (S450): 86–95. https://doi.org/10.1111/j.1651-2227.2006.tb02379.x.

O’Roak, B. J., H. A. Stessman, E. A. Boyle, K. T. Witherspoon, B. Martin, C. Lee, L. Vives, et al. 2014. “Recurrent de Novo Mutations Implicate Novel Genes Underlying Simplex Autism Risk.” Nature Communications 5 (November): 5595. https://doi.org/10.1038/ncomms6595.

O’Roak, B. J., Laura Vives, Santhosh Girirajan, Emre Karakoc, Niklas Krumm, Bradley P. Coe, Roie Levy, et al. 2012. “Sporadic Autism Exomes Reveal a Highly Interconnected Protein Network of de Novo Mutations.” Nature 485 (7397): 246–50. https://doi.org/10.1038/nature10989.

Pardiñas, Antonio F., Peter Holmans, Andrew J. Pocklington, Valentina Escott-Price, Stephan Ripke, Noa Carrera, Sophie E. Legge, et al. 2018. “Common Schizophrenia Alleles Are Enriched in Mutation-Intolerant Genes and in Regions under Strong Background Selection.” Nature Genetics 50 (3): 381–89. https://doi.org/10.1038/s41588-018-0059-2.

Parikshak, Neelroop N., Michael J. Gandal, and Daniel H. Geschwind. 2015. “Systems Biology and Gene Networks in Neurodevelopmental and Neurodegenerative Disorders.” Nature Reviews. Genetics 16 (8): 441–58. https://doi.org/10.1038/nrg3934.

Pinto, Dalila, Elsa Delaby, Daniele Merico, Mafalda Barbosa, Alison Merikangas, Lambertus Klei, Bhooma Thiruvahindrapuram, et al. 2014. “Convergence of Genes and Cellular Pathways Dysregulated in Autism Spectrum Disorders.” American Journal of Human Genetics 94 (5): 677–94. https://doi.org/10.1016/j.ajhg.2014.03.018.

Polioudakis, Damon, Luis de la Torre-Ubieta, Justin Langerman, Andrew G. Elkins, Xu Shi, Jason L. Stein, Celine K. Vuong, et al. 2019. “A Single Cell Transcriptomic Atlas of Human Neocortical Development during Mid-Gestation.” Neuron 103 (5): 785–801.e8. https://doi.org/10.1016/j.neuron.2019.06.011.

Poplin, Ryan, Valentin Ruano-Rubio, Mark A. DePristo, Tim J. Fennell, Mauricio O. Carneiro, Geraldine A. Van der Auwera, David E. Kling, et al. 2018. “Scaling Accurate Genetic Variant Discovery to Tens of Thousands of Samples.” https://doi.org/10.1101/201178.

Purcell, Shaun, Benjamin Neale, Kathe Todd-Brown, Lori Thomas, Manuel A. R. Ferreira, David Bender, Julian Maller, et al. 2007. “PLINK: A Tool Set for Whole-Genome Association and Population-Based Linkage Analyses.” American Journal of Human Genetics 81 (3): 559–75. https://doi.org/10.1086/519795.

Qiu, Anqi, Marcy Adler, Deana Crocetti, Michael I. Miller, and Stewart H. Mostofsky. 2010. “Basal Ganglia Shapes Predict Social, Communication, and Motor Dysfunctions in Boys with Autism Spectrum Disorder.” Journal of the American Academy of Child and Adolescent Psychiatry 49 (6): 539–51, 551.e1-4. https://doi.org/10.1016/j.jaac.2010.02.012.

Raven, John, and Jean Raven. 2003. “Raven Progressive Matrices.” In Handbook of Nonverbal Assessment, edited by R. Steve McCallum, 223–37. Boston, MA: Springer US. https://doi.org/10.1007/978-1-4615-0153-4_11.

Ronemus, Michael, Ivan Iossifov, Dan Levy, and Michael Wigler. 2014. “The Role of de Novo Mutations in the Genetics of Autism Spectrum Disorders.” Nature Reviews. Genetics 15 (2): 133–41. https://doi.org/10.1038/nrg3585.

Ruderfer, Douglas M., Stephan Ripke, Andrew McQuillin, James Boocock, Eli A. Stahl, Jennifer M. Whitehead Pavlides, Niamh Mullins, et al. 2018. “Genomic Dissection of Bipolar Disorder and Schizophrenia, Including 28 Subphenotypes.” Cell 173 (7): 1705–1715.e16. https://doi.org/10.1016/j.cell.2018.05.046.

Rutter, Michael, Ann Le Couteur, and Catherine Lord. 2013. ADI-R: Autism Diagnostic Interview Revised. Edited by Western Psychological Services. WPS edition. Los Angeles, CA: Western Psychological Services.

Ruzzo, Elizabeth K., Laura Pérez-Cano, Jae-Yoon Jung, Lee-Kai Wang, Dorna Kashef-Haghighi, Chris Hartl, Chanpreet Singh, et al. 2019. “Inherited and De Novo Genetic Risk for Autism Impacts Shared Networks.” Cell 178 (4): 850–866.e26. https://doi.org/10.1016/j.cell.2019.07.015.

Samocha, Kaitlin E., Jack A. Kosmicki, Konrad J. Karczewski, Anne H. O’Donnell-Luria, Emma Pierce-Hoffman, Daniel G. MacArthur, Benjamin M. Neale, and Mark J. Daly. 2017. “Regional Missense Constraint Improves Variant Deleteriousness Prediction,” June, 148353. https://doi.org/10.1101/148353.

Samocha, Kaitlin E., Elise B. Robinson, Stephan J. Sanders, Christine Stevens, Aniko Sabo, Lauren M. McGrath, Jack A. Kosmicki, et al. 2014. “A Framework for the Interpretation of de Novo Mutation in Human Disease.” Nature Genetics 46 (9): 944–50. https://doi.org/10.1038/ng.3050.

Sanders, Stephan J., Xin He, A. Jeremy Willsey, A. Gulhan Ercan-Sencicek, Kaitlin E. Samocha, A. Ercument Cicek, Michael T. Murtha, et al. 2015. “Insights into Autism Spectrum Disorder Genomic Architecture and Biology from 71 Risk Loci.” Neuron 87 (6): 1215– 33. https://doi.org/10.1016/j.neuron.2015.09.016.

Sandin, Sven, Paul Lichtenstein, Ralf Kuja-Halkola, Christina Hultman, Henrik Larsson, and Abraham Reichenberg. 2017. “The Heritability of Autism Spectrum Disorder.” JAMA 318 (12): 1182–84. https://doi.org/10.1001/jama.2017.12141.

Satija, Rahul, Jeffrey A. Farrell, David Gennert, Alexander F. Schier, and Aviv Regev. 2015. “Spatial Reconstruction of Single-Cell Gene Expression Data.” Nature Biotechnology 33 (5): 495–502. https://doi.org/10.1038/nbt.3192.

Satterstrom, F. Kyle, Jack A. Kosmicki, Jiebiao Wang, Michael S. Breen, Silvia De Rubeis, Joon-Yong An, Minshi Peng, et al. 2020. “Large-Scale Exome Sequencing Study Implicates Both Developmental and Functional Changes in the Neurobiology of Autism.” Cell 180 (3): 568–584.e23. https://doi.org/10.1016/j.cell.2019.12.036.

Satterstrom, F. Kyle, Raymond K. Walters, Tarjinder Singh, Emilie M. Wigdor, Francesco Lescai, Ditte Demontis, Jack A. Kosmicki, et al. 2019. “Autism Spectrum Disorder and Attention Deficit Hyperactivity Disorder Have a Similar Burden of Rare Protein-Truncating Variants.” Nature Neuroscience 22 (12): 1961–65. https://doi.org/10.1038/s41593-019-0527-8.

Sebat, Jonathan, B. Lakshmi, Dheeraj Malhotra, Jennifer Troge, Christa Lese-Martin, Tom Walsh, Boris Yamrom, et al. 2007. “Strong Association of de Novo Copy Number Mutations with Autism.” Science (New York, N.Y.) 316 (5823): 445–49. https://doi.org/10.1126/science.1138659.

Serdarevic, Fadila, Henning Tiemeier, Philip R. Jansen, Silvia Alemany, Yllza Xerxa, Alexander Neumann, Elise Robinson, Manon H. J. Hillegers, Frank C. Verhulst, and Akhgar Ghassabian. 2020. “Polygenic Risk Scores for Developmental Disorders, Neuromotor Functioning During Infancy, and Autistic Traits in Childhood.” Biological Psychiatry 87 (2): 132–38. https://doi.org/10.1016/j.biopsych.2019.06.006.

Simons Foundation Autism Research Initiative. 2020. “SFARI Gene 3.0.” SFARI Gene 3.0. September 16, 2020. https://gene.sfari.org/.

Singh, Tarjinder, James T. R. Walters, Mandy Johnstone, David Curtis, Jaana Suvisaari, Minna Torniainen, Elliott Rees, et al. 2017. “The Contribution of Rare Variants to Risk of Schizophrenia in Individuals with and without Intellectual Disability.” Nature Genetics 49 (8): 1167–73. https://doi.org/10.1038/ng.3903.

Skene, Nathan G., and Seth G. N. Grant. 2016. “Identification of Vulnerable Cell Types in Major Brain Disorders Using Single Cell Transcriptomes and Expression Weighted Cell Type Enrichment.” Frontiers in Neuroscience 10: 16. https://doi.org/10.3389/fnins.2016.00016.

Spielman, R. S., R. E. McGinnis, and W. J. Ewens. 1993. “Transmission Test for Linkage Disequilibrium: The Insulin Gene Region and Insulin-Dependent Diabetes Mellitus (IDDM).” American Journal of Human Genetics 52 (3): 506–16.

Starkstein, Sergio, Scott Gellar, Morgan Parlier, Leslie Payne, and Joseph Piven. 2015. “High Rates of Parkinsonism in Adults with Autism.” Journal of Neurodevelopmental Disorders 7 (1). https://doi.org/10.1186/s11689-015-9125-6.

Stein, Jason L., Luis de la Torre-Ubieta, Yuan Tian, Neelroop N. Parikshak, Israel A. Hernández, Maria C. Marchetto, Dylan K. Baker, et al. 2014. “A Quantitative Framework to Evaluate Modeling of Cortical Development by Neural Stem Cells.” Neuron 83 (1): 69–86. https://doi.org/10.1016/j.neuron.2014.05.035.

Stessman, Holly A. F., Bo Xiong, Bradley P. Coe, Tianyun Wang, Kendra Hoekzema, Michaela Fenckova, Malin Kvarnung, et al. 2017. “Targeted Sequencing Identifies 91 Neurodevelopmental-Disorder Risk Genes with Autism and Developmental-Disability Biases.” Nature Genetics 49 (4): 515–26. https://doi.org/10.1038/ng.3792.

Takahashi, Nagahide, Taeko Harada, Tomoko Nishimura, Akemi Okumura, Damee Choi, Toshiki Iwabuchi, Hitoshi Kuwabara, et al. 2020. “Association of Genetic Risks With Autism Spectrum Disorder and Early Neurodevelopmental Delays Among Children Without Intellectual Disability.” JAMA Network Open 3 (2): e1921644. https://doi.org/10.1001/jamanetworkopen.2019.21644.

Torre-Ubieta, Luis de la, Hyejung Won, Jason L. Stein, and Daniel H. Geschwind. 2016. “Advancing the Understanding of Autism Disease Mechanisms through Genetics.” Nature Medicine 22 (4): 345–61. https://doi.org/10.1038/nm.4071.

Turner, Tychele N., Bradley P. Coe, Diane E. Dickel, Kendra Hoekzema, Bradley J. Nelson, Michael C. Zody, Zev N. Kronenberg, et al. 2017. “Genomic Patterns of De Novo Mutation in Simplex Autism.” Cell 171 (3): 710–722.e12. https://doi.org/10.1016/j.cell.2017.08.047.

Turner, Tychele N., Fereydoun Hormozdiari, Michael H. Duyzend, Sarah A. McClymont, Paul W. Hook, Ivan Iossifov, Archana Raja, et al. 2016. “Genome Sequencing of Autism-Affected Families Reveals Disruption of Putative Noncoding Regulatory DNA.” American Journal of Human Genetics 98 (1): 58–74. https://doi.org/10.1016/j.ajhg.2015.11.023.

Van der Auwera, Geraldine A., Mauricio O. Carneiro, Christopher Hartl, Ryan Poplin, Guillermo Del Angel, Ami Levy-Moonshine, Tadeusz Jordan, et al. 2013. “From FastQ Data to High Confidence Variant Calls: The Genome Analysis Toolkit Best Practices Pipeline.” Current Protocols in Bioinformatics 43: 11.10.1–11.10.33. https://doi.org/10.1002/0471250953.bi1110s43.

Van der Auwera, Geraldine A., and Brian D. O’Connor. 2020. Genomics in the Cloud: Using Docker, GATK, and WDL in Terra. O’Reilly Media.

Veatch, Olivia J, Brendan T Keenan, Philip R Gehrman, Beth A Malow, and Allan I Pack. 2017. “Pleiotropic Genetic Effects Influencing Sleep and Neurological Disorders.” The Lancet. Neurology 16 (2): 158–70. https://doi.org/10.1016/S1474-4422(16)30339-8.

Vilhjálmsson, Bjarni J., Jian Yang, Hilary K. Finucane, Alexander Gusev, Sara Lindström, Stephan Ripke, Giulio Genovese, et al. 2015. “Modeling Linkage Disequilibrium Increases Accuracy of Polygenic Risk Scores.” The American Journal of Human Genetics 97 (4): 576–92. https://doi.org/10.1016/j.ajhg.2015.09.001.

Vorontsov, Ilya E., Alla D. Fedorova, Ivan S. Yevshin, Ruslan N. Sharipov, Fedor A. Kolpakov, Vsevolod J. Makeev, and Ivan V. Kulakovskiy. 2018. “Genome-Wide Map of Human and Mouse Transcription Factor Binding Sites Aggregated from ChIP-Seq Data.” BMC Research Notes 11 (1): 756. https://doi.org/10.1186/s13104-018-3856-x.

Wang, Jing, Leon Raskin, David C. Samuels, Yu Shyr, and Yan Guo. 2015. “Genome Measures Used for Quality Control Are Dependent on Gene Function and Ancestry.” Bioinformatics 31 (3): 318–23. https://doi.org/10.1093/bioinformatics/btu668.

Wang, Kai, Mingyao Li, and Hakon Hakonarson. 2010. “ANNOVAR: Functional Annotation of Genetic Variants from High-Throughput Sequencing Data.” Nucleic Acids Research 38 (16): e164. https://doi.org/10.1093/nar/gkq603.

Warrier, Varun, Xinhe Zhang, Patrick Reed, Alexandra Havdahl, Tyler M. Moore, Freddy Cliquet, Claire S. Leblond, et al. 2021. “Genetic Correlates of Phenotypic Heterogeneity in Autism.” https://doi.org/10.1101/2020.07.21.20159228.

Weiner, Daniel J., Emilie M. Wigdor, Stephan Ripke, Raymond K. Walters, Jack A. Kosmicki, Jakob Grove, Kaitlin E. Samocha, et al. 2017. “Polygenic Transmission Disequilibrium Confirms That Common and Rare Variation Act Additively to Create Risk for Autism Spectrum Disorders.” Nature Genetics 49 (7): 978–85. https://doi.org/10.1038/ng.3863.

Werling, Donna M., Harrison Brand, Joon-Yong An, Matthew R. Stone, Lingxue Zhu, Joseph T. Glessner, Ryan L. Collins, et al. 2018. “An Analytical Framework for Whole-Genome Sequence Association Studies and Its Implications for Autism Spectrum Disorder.” Nature Genetics 50 (5): 727–36. https://doi.org/10.1038/s41588-018-0107-y.

Werling, Donna M., and Daniel H. Geschwind. 2013. “Sex Differences in Autism Spectrum Disorders.” Current Opinion in Neurology 26 (2): 146–53. https://doi.org/10.1097/WCO.0b013e32835ee548.

Wickham, Hadley. 2009. Ggplot2: Elegant Graphics for Data Analysis. Use R! New York: Springer-Verlag. https://doi.org/10.1007/978-0-387-98141-3.

Xu, Danqing, Chen Wang, Krzysztof Kiryluk, Joseph D. Buxbaum, and Iuliana Ionita-Laza. 2020. “Co-Localization between Sequence Constraint and Epigenomic Information Improves Interpretation of Whole-Genome Sequencing Data.” American Journal of Human Genetics 106 (4): 513–24. https://doi.org/10.1016/j.ajhg.2020.03.003.

Yu, Guangchuang, Li-Gen Wang, Yanyan Han, and Qing-Yu He. 2012. “ClusterProfiler: An R Package for Comparing Biological Themes among Gene Clusters.” Omics: A Journal of Integrative Biology 16 (5): 284–87. https://doi.org/10.1089/omi.2011.0118.

Yuen, Ryan KC, Daniele Merico, Matt Bookman, Jennifer L Howe, Bhooma Thiruvahindrapuram, Rohan V Patel, Joe Whitney, et al. 2017. “Whole Genome Sequencing Resource Identifies 18 New Candidate Genes for Autism Spectrum Disorder.” Nature Neuroscience 20 (4): 602–11. https://doi.org/10.1038/nn.4524.

Yuen, Ryan KC, Bhooma Thiruvahindrapuram, Daniele Merico, Susan Walker, Kristiina Tammimies, Ny Hoang, Christina Chrysler, et al. 2015. “Whole-Genome Sequencing of Quartet Families with Autism Spectrum Disorder.” Nature Medicine 21 (2): 185–91. https://doi.org/10.1038/nm.3792.

Zhou, Jian, Christopher Y. Park, Chandra L. Theesfeld, Aaron K. Wong, Yuan Yuan, Claudia Scheckel, John J. Fak, et al. 2019. “Whole-Genome Deep-Learning Analysis Identifies Contribution of Noncoding Mutations to Autism Risk.” Nature Genetics 51 (6): 973–80. https://doi.org/10.1038/s41588-019-0420-0.

Zook, Justin M., Brad Chapman, Jason Wang, David Mittelman, Oliver Hofmann, Winston Hide, and Marc Salit. 2014. “Integrating Human Sequence Data Sets Provides a Resource of Benchmark SNP and Indel Genotype Calls.” Nature Biotechnology 32 (3): 246–51. https://doi.org/10.1038/nbt.2835.

